# Genetic underpinnings of the transition from alcohol consumption to alcohol use disorder: shared and unique genetic architectures in a cross-ancestry sample

**DOI:** 10.1101/2021.09.08.21263302

**Authors:** Rachel L. Kember, Rachel Vickers-Smith, Hang Zhou, Heng Xu, Cecilia Dao, Amy C. Justice, Joel Gelernter, Marijana Vujkovic, Henry R. Kranzler

## Abstract

Recent GWAS of alcohol-related traits have uncovered key differences in the underlying genetic architectures of alcohol consumption and alcohol use disorder (AUD), with the two traits having opposite genetic correlations with psychiatric disorders. Understanding the genetic factors that underlie the transition from heavy drinking to AUD has important theoretical and clinical implications. We utilized longitudinal data from the cross-ancestry Million Veteran Program sample to identify 1) novel loci associated with AUD and alcohol consumption [measured by the Alcohol Use Disorders Identification Test-Consumption (AUDIT-C)] and 2) genetic variants with direct effects on AUD not mediated through alcohol consumption. We identified 26 loci associated with AUD, including 5 ancestry-specific and 6 novel loci and 22 loci associated with AUDIT-C, including 3 ancestry-specific and 8 novel loci. In secondary GWAS that excluded individuals who report abstinence, we identify 7 additional loci for AUD and 8 additional loci for AUDIT-C. We demonstrate that, although the heterogeneity of the abstinent group biases the GWAS findings, unique variance between alcohol consumption and disorder remains after the group is excluded. Finally, using mediation analysis, we identified a set of variants with effects on AUD that are not mediated through alcohol consumption. The distinct genetic architectures of alcohol consumption and AUD suggest different biological contributions to the traits. Genetic variants with direct effects on AUD are potentially relevant to understanding the transition from heavy alcohol consumption to AUD and targets for translational prevention and treatment efforts.

## Introduction

Heavy alcohol use is common in the United States and associated with multiple adverse consequences(1). Regular heavy drinking is the major risk factor for alcohol use disorder (AUD), a problematic pattern of alcohol use accompanied by clinically significant impairment or distress(2). Recent genome-wide association studies (GWAS) of alcohol traits have identified more than 30 genome-wide significant (GWS) variants contributing to the risk of AUD, alcohol dependence (AD), or problematic alcohol use (PAU; which comprises AUD, AD, and a trait defined by alcohol-related problems)(3–5); and more than 100 GWS variants have been identified as contributing to alcohol consumption measures [e.g., AUDIT-C score(6), a measure of alcohol consumption derived from the first 3 items of the 10-item Alcohol Use Disorders Identification Test (AUDIT)(7)](4,8,9). Nearly all GWAS loci have been identified in European ancestry individuals, largely due to the much smaller numbers of individuals from other population groups for which comparable data are available.

Although the genetic variation contributing to alcohol consumption partially overlaps with that of AUD, multiple variants are associated with one, but not the other, trait(10). Estimates of the genetic correlation between alcohol consumption and either AUD or PAU are moderate(4,5), suggesting that despite alcohol use being necessary for dependence, there is unique genetic liability to each trait. Furthermore, genetic correlations and phenome-wide association studies (PheWAS) of genetic liability for AUD and alcohol consumption show differential patterns of association with physical and mental health(4,5,8).

The genetic divergence between alcohol consumption and AUD may result from confounding, or it may indicate a true difference in the biology underlying each trait. Potential sources of bias include confounding of the frequency of alcohol consumption by socioeconomic status(11), and the heterogeneity of participants reporting alcohol abstinence(12). However, differences in genetic liability between alcohol consumption and AUD are observed even after these confounders are accounted for. Thus, there may exist a latent factor that underlies psychopathology and substance use *disorders*, but not substance *use*(13). As AUD is defined by several diagnostic criteria not directly related to the frequency or quantity of alcohol consumption, it is possible for individuals with the same level of alcohol consumption to receive differential diagnoses for AUD. Consequently, understanding the factors underlying AUD, separate from those that influence normative alcohol consumption, has important theoretical and clinical implications.

We sought to identify genetic variants that contribute uniquely to the development of AUD by conducting multiple cross-ancestry GWASs in the Million Veteran Program (MVP)(14). To examine the transition from alcohol consumption to AUD, we conducted analyses to identify genetic variants that are not mediated through alcohol consumption, but rather directly affect the risk of AUD.

## Methods

### Overview of analyses

This research was conducted with three aims in mind. First, we sought to identify novel loci associated with AUD and alcohol consumption by performing separate ancestry-specific primary GWAS of AUD and AUDIT-C, followed by cross-ancestry meta-analyses. Second, we evaluated the mediating effect of alcohol consumption on genetic risk for AUD by conducting GWAS in which we restricted AUD cases to those with an AUDIT-C score that pre-dated their receiving an AUD diagnosis and removed abstainers from cases and controls. Third, in secondary analyses, we elucidated the impact of phenotypic variation on gene discovery by performing an additional GWAS using a less stringent electronic health record (EHR)-defined definition of AUD. Supplemental Figure 1 provides an overview of the phenotype definitions and sample counts for each analysis.

### Million Veteran Program cohort and phenotype construction

The MVP is a large observational cohort study and biobank developed by the Department of Veterans Affairs (VA)(14). AUD diagnostic codes based on the International Classification of Diseases, Ninth Revision (ICD-9) and Tenth Revision (ICD-10) and AUDIT-C scores were obtained from the VA EHR. The AUDIT-C(6) comprises the first three questions of the 10-item AUDIT, a valid, reliable instrument used to screen for individuals with hazardous or harmful drinking(7). We use the maximum AUDIT-C score (“AUDIT-C”) to approximate a trait, rather than state, alcohol consumption measure. The dataset comprised all MVP participants with at least one AUDIT-C measure between 2007 to 2018. In all analyses, age at the time of the maximum AUDIT-C score was used as a covariate in GWAS. Further details on the phenotypes used for each analysis are described in the Supplemental Methods.

### Genome-wide association analysis

For ancestry-specific analyses, we tested imputed SNPs that passed quality control (see Supplemental Methods) for association with 5 traits: 1) AUD (Stringent) (primary analysis), 2) AUDIT-C (primary analysis), AUD only among AUDIT-C>0 (mediation analysis) and AUDIT-C only among AUDIT-C>0 (mediation analysis), and AUD (Less stringent) (secondary analysis). The analyses used logistic or linear regression models as appropriate, in PLINK 1.9(15). Covariates included age at maximum AUDIT-C, sex, and the first 10 ancestral principal components (PCs). Cross-ancestry meta-analyses were performed in METAL(16) using the sample-size weighted method. Within each region, independent variants were identified by conditional analyses using GCTA-COJO(17) (see Supplemental Methods for additional details).

### Gene-based analyses, heritability analyses and genetic correlation

Gene-based association analyses for the primary AUD and AUDIT-C GWAS were performed using MAGMA(18) implemented in FUMA(19). Default settings were used to map input SNPs to 19,082 protein-coding genes. Genome-wide significance was defined at P = 0.05/19,082 = 2.62×10^−6^. SNP-based heritability (h^2^) for all phenotypes was estimated using linkage disequilibrium score regression (LDSC(20)). Genetic correlation analyses were performed using LDSC(20) and POPCORN(21) (see Supplemental Methods).

### Mediation

To identify a set of variants for mediation analyses, we selected all variants associated with either AUD (Stringent) or AUDIT-C at a suggestive p-value threshold of 1×10^−5^ in the cross-ancestry meta-analyses. Among variants present in all 3 ancestries, 3,092 were selected for AUD and 3,019 for AUDIT-C. We performed LD-clumping using a range of 3000 kb and r^2^ > 0.1 to create a set of 215 variants for AUD and 208 variants for AUDIT-C, yielding a total set of 372 unique variants associated with either AUD, AUDIT-C, or both.

The variant set was tested for association with a) AUD only among AUDIT-C>0; b) AUDIT-C>0; and c) AUD only among AUDIT-C>0 with AUDIT-C as a covariate. All analyses included age, sex and the first 10 PCs as covariates. The direct effect of the variant on AUD was estimated from the association of the variant with AUD with AUDIT-C as a covariate. The mediated (indirect) effect of the variant on AUD through AUDIT-C was calculated using the regression method(22) from summary-level data for the association of the variant with the outcome (i.e., AUD) including the mediator (i.e., covarying for AUDIT-C) and the mediator without the outcome (i.e., AUDIT-C). Analyses were conducted separately for each ancestry before calculating cross-ancestry meta-analyses in METAL. Variants that passed a Bonferroni-corrected p-value threshold of 1.34×10^−4^ (0.05/372) in the AUD (AUDIT-C>0) analysis (i.e., were associated with the outcome) were explored to partition the total effect into a direct effect and an indirect effect.

## Results

We included 409,630 MVP participants from three ancestral groups: European Americans (EAs) (N=296,989), African Americans (AAs) (N=80,764) and Hispanic Americans (HAs) (N=31,877) (Supplemental Table 1). Most of the population was male (91.5%). Nearly one-quarter (24.7%) of participants had at least one ICD-9/10 code for AUD (i.e., AUD [less stringent] EAs: 21.0%, AAs: 35.2%, HAs: 29.2%) and 20.8% had at least one inpatient or two outpatient codes for AUD (i.e., AUD [Stringent] EAs: 18.0%, AAs: 31.0%, HAs: 24.6%). The average number of annual AUDIT-C screenings was 8.1 (SD=3.8; EAs=8.1, SD=3.7; AAs=8.5, SD=3.9; HAs=7.6, SD=3.7). Among individuals with an AUD diagnosis, the average number of AUDIT-C screenings that pre-dated the diagnosis was 3.3 (SD=2.8; EAs=3.4, SD=2.9; AAs=3.2, SD=2.7; HAs=3.1, SD=2.6). The average maximum AUDIT-C score for all individuals was 3.0, with an average age at maximum AUDIT-C of 59.8 years. Maximum AUDIT-C mean scores were higher in AUD cases (5.6) than controls (2.2) (Supplemental Figure 2).

### Identification of cross-ancestry and ancestry-specific novel loci for AUD and AUDIT-C

We performed ancestry-specific genome-wide analyses for AUD and AUDIT-C, followed by cross-ancestry meta-analyses (Supplemental Figure 3). In the cross-ancestry meta-analysis for AUD, 84 lead variants (mapping to 21 loci; Figure 1, Supplemental Table 2) were GWS (P<5×10^−8^). After conditioning on the lead variants within each locus, 26 variants were independently associated with AUD, of which 4 are novel (no previous reported associations with alcohol-related phenotypes within 1 Mb, Supplemental Table 4), including two that are intronic (rs11681373 in *ZNF804A* and rs3828783 in *MLN*) and two intergenic (rs138084484 near *NICN1* and rs565298761 near *MIR5694*). Supplemental Figure 4 and Supplemental Table 3 show the 27 lead variants (in 14 loci) in EAs, 5 lead variants (in 3 loci) in AAs, and 4 lead variants (in 1 locus) in HAs. Of the lead variants identified in EAs, 4 variants in 3 loci were GWS in the ancestry-specific analysis only (Supplemental Figure 5), of which one is novel (rs34517574 near *MIR129-2*, Supplemental Table 4). All EA ancestry-specific variants were polymorphic in all populations (minor allele frequency [MAF] >10%), with the exception of rs34517574, which was not present in the AA sample due to low imputation quality. In AAs, 2 lead variants in 2 loci were ancestry-specific (Supplemental Figure 6), of which one is novel (rs7701111 in an intron of *IL7R*, Supplemental Table 4). Although one of these variants was rare in EAs and HAs (rs73792114; MAF: AA=5.9%, EA=0.04%, HA=0.08%), the novel variant was polymorphic in all populations (rs7701111; MAF: AA=20.4%, EA=32.7%, HA=42.1%).

**Figure 1:**
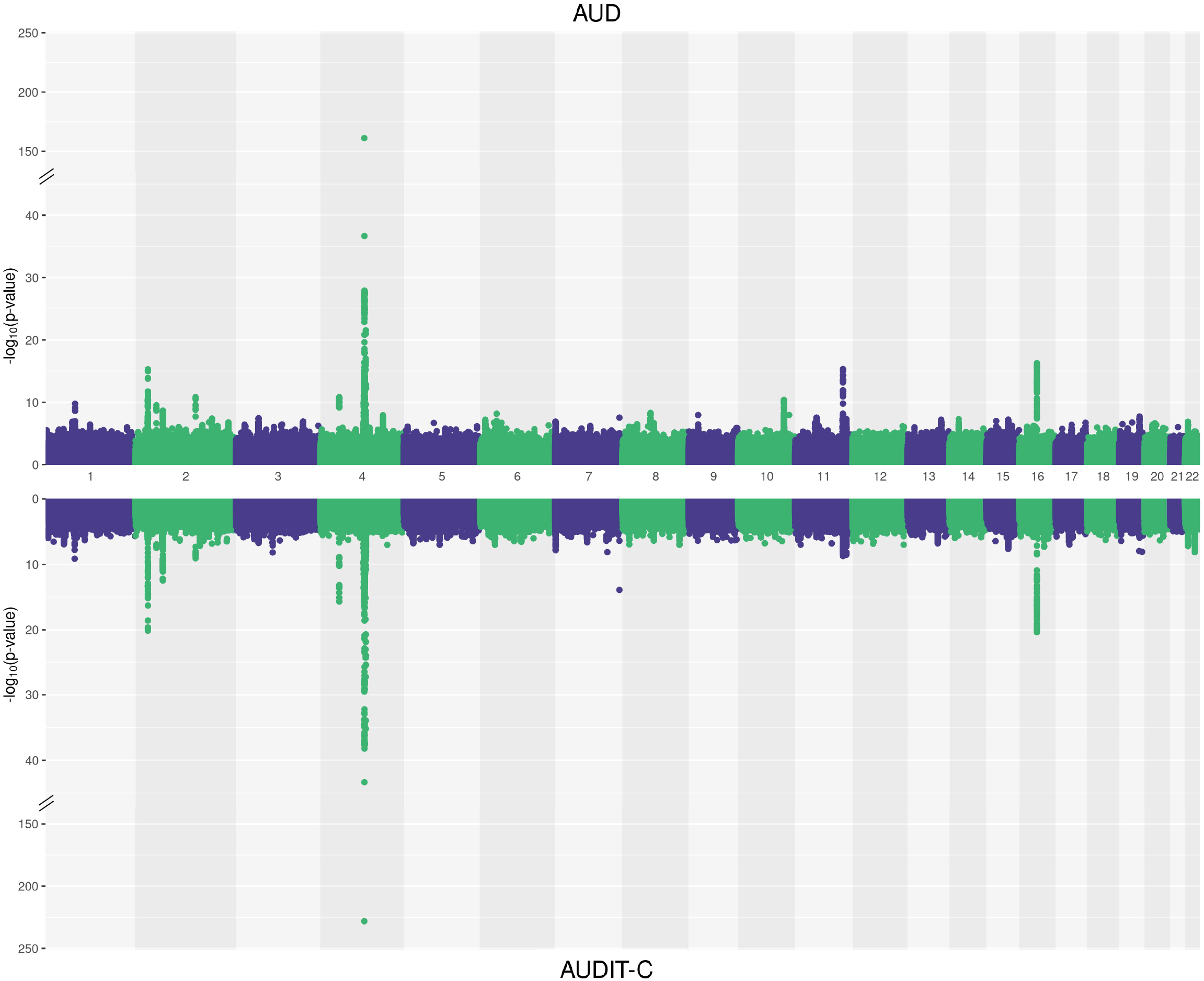
Manhattan plot for cross-ancestry meta-analysis of AUD (top, N case=85,391, N control=308,488) and AUDIT-C (bottom, N=409,630). Sample-size-weighted meta-analyses were performed using METAL.

In the cross-ancestry meta-analysis for AUDIT-C, 87 lead variants (mapping to 19 loci; Figure 1, Supplemental Table 5) were identified. Conditional analysis yielded 24 independent variants, of which 6 are novel (Supplemental Table 7). Two of the novel variants are intronic (rs34305371 in *NEGR1* and rs307914 in *CACNG7*) and four intergenic (rs62260887 near *GBE1*, rs12702456 near *LOC101927021*, rs144228648 near *POT1*, and rs8041398 near *HMG20A*). In ancestry-specific analyses (Supplemental Figure 7 and Supplemental Table 6), 37 variants (in 14 loci) were GWS in EAs, 3 variants (in 2 loci) were GWS in AAs, and 4 variants (in 1 locus) were GWS in HAs. In EAs, 2 variants in 2 loci were ancestry-specific, of which one was novel and polymorphic in all populations (Supplemental Figure 8, Supplemental Table 7; rs13389219 near *COBLL1*). In AAs, there was one ancestry-specific variant, which was novel (Supplemental Figure 9, Supplemental Table 7; rs150461125 near *OLFM1*), though rare in AAs, almost monomorphic in EAs, and absent in HAs (MAF: AA=0.002%, EA<0.001%).

We conducted three additional GWAS: 1) AUD in individuals with AUDIT-C>0, 2) AUDIT-C in individuals with AUDIT-C>0, and 3) AUD using a less stringent diagnosis (Supplemental Figures 3 and 10-12, Supplemental Tables 8-13). In all analyses, correlations between effect sizes for GWS loci identified in the primary GWAS and the secondary GWAS were high (r^2^=0.99).

**Figure 3:**
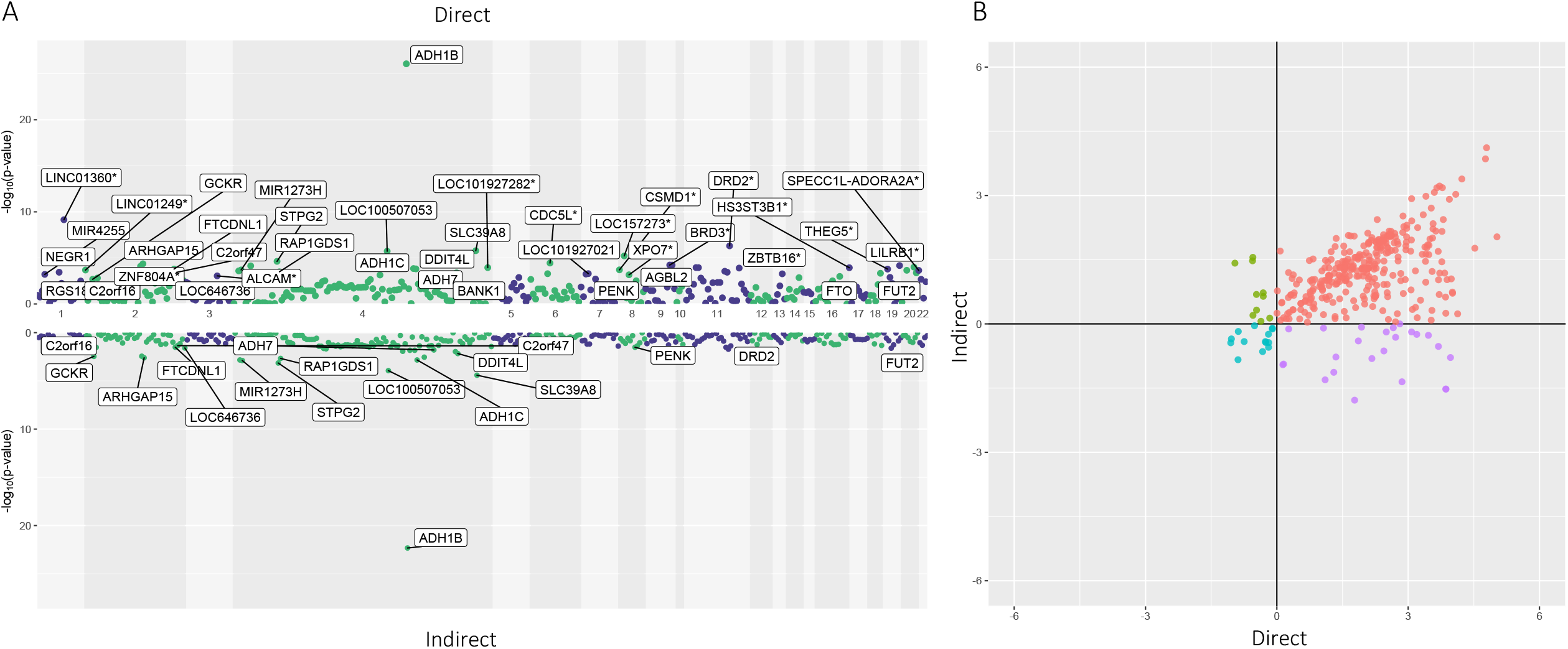
Mediation analysis of variants associated with AUD. A: Manhattan plots showing direct effects of variants on AUD (top) and indirect effects mediated by AUDIT-C (bottom). Variants significantly associated with AUD at a Bonferroni corrected p-value, and then subsequently having a direct and/or indirect effect, are labelled. For ease of visualization, only the most significant SNP in or near each gene is labelled. Variants where more than two thirds of the effect is direct are labelled with an asterisk. B: Effect sizes (Z-scores) for each variant. Direct effects are shown on the x-axis and indirect effects are shown on the y-axis. The majority of variants have the same direction of effect for both direct and indirect effects.

### Gene-based analyses

Gene-based association analyses for AUD identified 58 GWS (p⍰<⍰2.69×10^™6^) genes in EAs, 10 in AAs, and 7 in HAs (Supplemental Figure 13). Genes not identified in the SNP-based analyses of AUD (EA: N=47, AA: N=8, HA: N=4) include *ADH4*, which was GWS in both EAs and AAs, *ADH5*, which was GWS in AAs, and *ADH7*, which was GWS in HAs. Gene-based association analyses for AUDIT-C identified 45 GWS genes in EAs, 7 in AAs, and 6 in HAs (Supplemental Figure 14). In EAs, 38 of these were not identified in SNP-based analyses, including *CACNA1C*; 6 in AAs, including *METAP1*; and 3 in HAs, including *TRMT10A*.

### Heritability and genetic correlation of AUD and AUDIT-C phenotypes

SNP-based heritability (h^2^_SNP_) for the five related GWAS traits was estimated using LDSC for both EAs and AAs (Figure 2a). Heritability values were higher for AAs than EAs and increased with the specificity of the phenotype. Pairwise genetic correlations (r_g_) between the five GWAS traits were high within both EAs and AAs (r_g_>0.7; Supplemental Figure 15). Correlations were strongest within the AUD and AUDIT-C phenotypes, and weaker between the two kinds of traits. Genetic correlations between EAs and AAs within traits were estimated using POPCORN (Supplemental Table 14).

**Figure 2:**
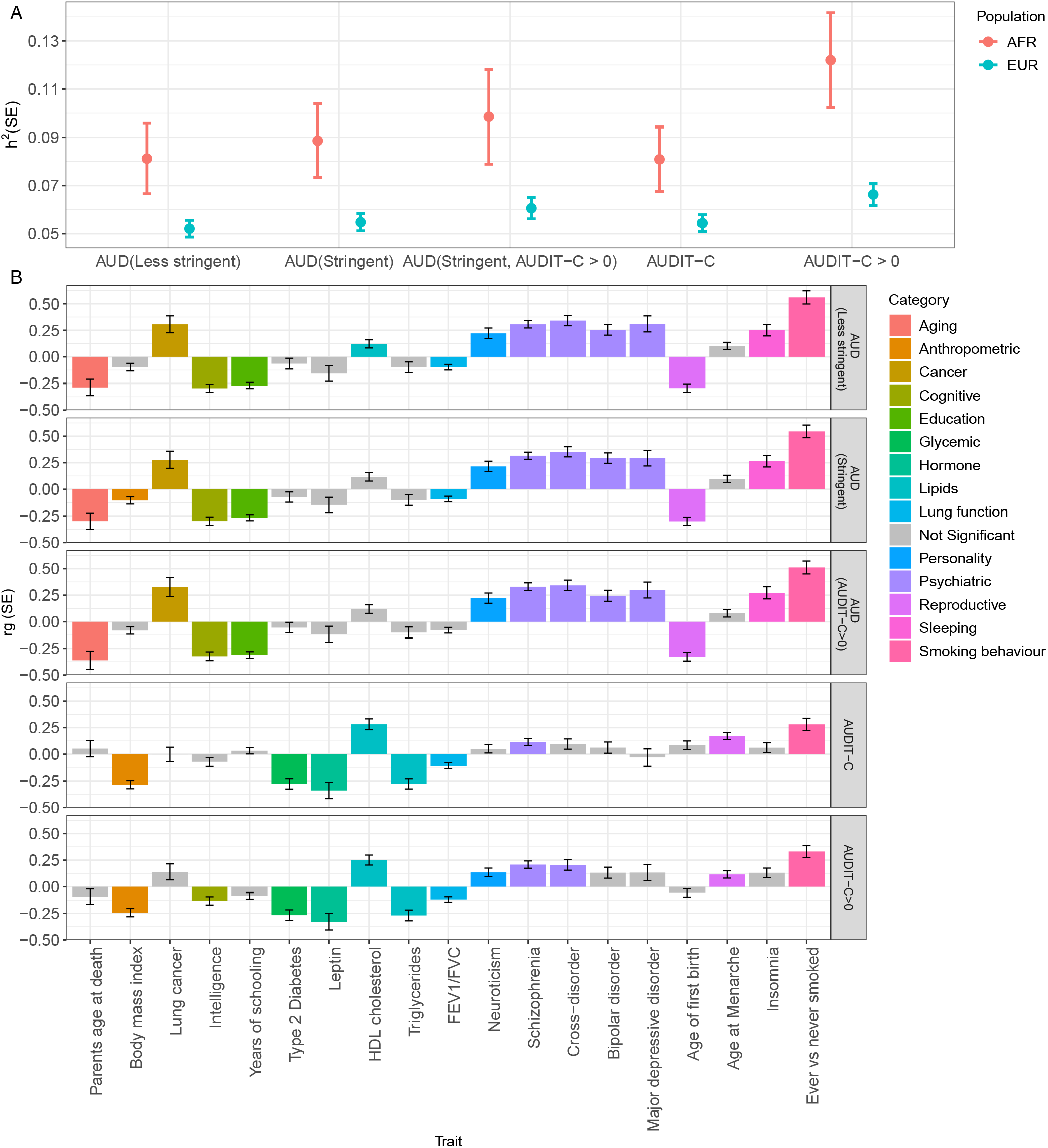
Heritability and genetic correlation analyses for AUD (less stringent), AUD, AUD (AUDIT-C>0), AUDIT-C, and AUDIT-C>0. A: Heritability calculated for EAs and AAs using LDSC. B: Genetic correlations with other traits in EAs calculated using LDSC. Non-grey bars are significant at a Bonferroni-corrected p-value (p<1.95×10^−3^).

### Genetic correlations with other traits

The genetic correlations between the five GWAS traits and other traits or diseases were estimated in the EA population using LDSC (Figure 2b, Supplemental Table 15). As in our previous work(4), the magnitude and direction of genetic correlations differed substantially between AUD and AUDIT-C. AUD was correlated with 22 traits, including positive correlations with several psychiatric disorders, insomnia, and neuroticism, and negative correlations with years of schooling and age at first birth. In contrast, AUDIT-C was positively genetically correlated with HDL cholesterol and lung function and negatively correlated with anthropometric traits and Type 2 diabetes. Traits significantly genetically correlated with AUD (AUDIT-C>0) and AUD (less stringent) were generally the same as those correlated with AUD. Many of the traits that were significantly genetically correlated with AUDIT-C were also correlated with AUDIT-C>0 in the same direction. However, AUDIT-C>0 was also significantly positively correlated with psychiatric traits.

### Mediation

In mediation analysis we identified total effects of variants on AUD and AUDIT-C, and direct and indirect effects of variants on AUD (Figure 3 and Supplemental Tables 16-19). In the cross-ancestry meta-analysis, 54 variants were associated with AUD at a Bonferroni-corrected p-value threshold of 1.34×10^−4^. For 17 of these (31%) the direct effect on AUD accounted for more than two-thirds of the total effect, indicating that AUD susceptibility at these loci was not predominantly driven by a genetic predisposition for alcohol consumption.

Most variants that showed evidence for mediation had a concordant directionality for the direct and indirect effects (Figure 3B). The strongest association with both AUD and AUDIT-C was rs1229984 in *ADH1B* (Figure 3A, Supplemental Table 16). In the mediation analysis, this SNP showed both a strong direct and a strong indirect effect on AUD. Other variants with both strong direct and indirect effects on AUD included rs13107325 in *SLC39A8*, rs1154431 in *LOC100507053*, rs138081390 in *STPG2* and rs3216150 in *ADH1C*. Variants in *GCKR* (rs11336847) and *FTO* (rs1421085, rs55872725) had almost equal direct and indirect effect sizes.

Of the 17 variants with predominantly direct effects on AUD, the strongest effects were for rs7517355 in *LINC01360*, rs61687445 near *DRD2*, rs73201625 near *LOC157273*, rs114569212 near *CDC5L*, two variants near *BRD3* (rs146671954, rs146281019), and rs61739176 near *LILRB1*. Variants near *LPHN3* (rs115663100) and *TMX2* (rs79260745, rs7948571), and in an intron of *SLC9A8* (rs76728843) had strong direct effects on AUD but an indirect effect in the opposite direction, reducing their overall association with AUD.

## Discussion

Here, we performed cross-ancestry GWAS of AUD and AUDIT-C in 409,630 individuals, including the largest sample of non-European-ancestry individuals for these traits to date (AA: N=80,764, HA: N=31,877). These populations were 40-50% larger than in our previous GWAS for AUDIT-C(4) and AUD,(4) which allowed us to identify novel loci in both the cross-ancestry meta-analysis and the ancestry-specific analyses. We conducted mediation analysis to establish a set of variants directly associated with AUD, independent of alcohol consumption. We propose that these variants are high-priority candidates for *in-vitro* study to understand the biology of the transition from alcohol use to the development of AUD, as they could help account for why individuals with the same level of alcohol consumption differ in their risk to develop AUD.

We identified 26 loci associated with AUD, including 4 novel loci in the cross-ancestry meta-analysis and 2 novel loci in ancestry-specific analyses. Many of the loci, though novel for alcohol-related traits, have been associated with psychiatric disorders or other substance use or disorders. One novel intronic variant (rs11681373) in *ZNF804A* (Zinc Finger Protein 804A) is located in intron 2, close to a variant previously associated with increased susceptibility to a broad psychosis trait (including schizophrenia and bipolar disorder) and heroin addiction(23). The second novel intronic variant (rs3828783) is located in *MLN* (Motilin), a gene previously associated with smoking(9), and depression(24). A novel intergenic variant (rs138084484) located upstream of *NICN1* (Nicolin 1), which has been associated with depression(25) and intelligence(26), is a known eQTL for multiple other genes(27). The novel AA-specific variant is located in the intron of *IL7R* (Interleukin 7 Receptor), which plays a critical role in the development of immune cells(28). Chronic alcohol consumption is known to impair immune response functions, and concentrations of IL7 are altered during detoxification(29).

Twenty-two loci were associated with AUDIT-C, including 6 novel loci in the cross-ancestry meta-analysis and 2 novel variants in the ancestry-specific analyses. No prior associations with psychiatric disorders have been identified for these novel loci, consistent with the observed divergence in the genetic architectures of the AUDIT-C and AUD(10). Both of the novel intronic variants are located in genes with prior associations with intelligence and educational attainment (rs34305371 in *NEGR1* [Neuronal Growth Regulator 1] and rs307914 in *CACNG7* [Calcium Voltage-Gated Channel Auxiliary Subunit Gamma 7])(26,30). The novel EA-specific variant rs13389219 near *COBLL1* has been associated with multiple phenotypes including Type 2 diabetes(31), whereas variants located nearby have shown associations with HDL cholesterol and triglyceride measures in current drinkers(32). The novel AA-specific variant rs150461125 is located near *OLFM1*, a gene associated with smoking behavior(9).

We refined both the AUD and AUDIT-C phenotypes by removing individuals who never report alcohol consumption. In the UK Biobank and the Veterans Aging Cohort Study, where participants’ specific reasons for discontinuing drinking was available, we found that this group is heterogeneous, consisting of both lifelong abstainers and former drinkers(12). Removing them from the analyses did not increase the number of GWS loci, likely in part because they make up almost a quarter of our original sample. However, we observed an increase in the SNP heritability for both AUD and AUDIT-C following their removal. Furthermore, in genetic correlation analyses with other traits, the removal of these individuals revealed positive correlations of AUDIT-C with psychiatric traits and a negative correlation with intelligence.

Evaluating the mediating effect of AUDIT-C on the genetic risk of AUD identified variants in three alcohol metabolizing genes (alcohol dehydrogenase 1B [*ADH1B*], 1C [*ADH1C*] and 7 [*ADH7*]) with both direct and indirect effects on AUD risk. Variants in these genes alter the metabolic rate of alcohol and consistently are associated with protective effects against excessive alcohol consumption, thereby reducing AUD risk(33). The direct (non-mediated) effect of these variants on AUD could result from their effects on alcohol consumption earlier in life, prior to the measurement of AUDIT-C, or convey risk of AUD via an independent pathway.

The variant with the second strongest direct and indirect effects on AUD was in *SLC39A8* (solute carrier family 39 member 8; rs13107325), which has highly pleiotropic effects, having been associated with over 60 traits(34), including anthropometric traits such as BMI(35), and psychiatric traits such as schizophrenia(36). Thus, this variant may affect risk for AUD both via alcohol consumption and via effects on the psychopathological dimension of addiction.

We identified 17 variants for which greater than two-thirds of the effect on AUD risk was direct, i.e., not mediated through alcohol consumption. We hypothesize that these variants contribute to an addictive dimension of AUD that is not directly influenced by levels of consumption. Of these, 4 variants were in or near genes with prior associations with schizophrenia(36) (*LINC01360* [Long Intergenic Non-Protein Coding RNA 1360], *DRD2* [Dopamine Receptor D2], *CSMD1* [CUB And Sushi Multiple Domains 1], and *ZNF804A* [Zinc Finger Protein 804A]), three of which (*DRD2, CSMD1* and *ZNF804A*) are mostly expressed in brain(27). Other variants were in or near genes previously associated with cortical surface area (*ALCAM* [Activated Leukocyte Cell Adhesion Molecule])(37), chronotype (*LINC01249* [Long Intergenic Non-Protein Coding RNA 1249])(38), lung function (*CDC5L* [Cell Division Cycle 5 Like])(39), and caffeine consumption (*SPECC1L-ADORA2A* [Readthrough (NMD Candidate)])(40). These variants warrant further examination as potential contributors to the transition from heavy drinking to AUD.

Our study has limitations. Our measure of alcohol consumption, the AUDIT-C, was self-reported and subject to recall bias. Further, we systematically captured consumption reported in the VA health system only since 2007. Therefore, if an individual’s maximum alcohol consumption occurred prior to this date this measure may not be accurate. Furthermore, the AUDIT-C measure is made up of three items that capture quantity and frequency of drinking and heavy drinking. Different responses to these items could result in the same overall score, and yet each item may be differentially associated with AUD. Finally, the MVP sample is mostly male (>90%) and we therefore could not perform sex-specific analyses. As AUDIT-C scores vary by sex, this will be an important direction for future studies.

In conclusion, our study identifies novel associations for AUD and AUDIT-C, including novel ancestry-specific associations. We address some of the differences between the genetic architectures of AUD and AUDIT-C by reducing heterogeneity in the dataset, but also demonstrate that there is unique variance that remains after refinement of these traits. Finally, we identify a set of variants with direct effects on AUD that are not mediated through alcohol consumption. We propose these as targets for research aimed at understanding the transition from heavy alcohol consumption to AUD, elucidation of the biology of which could inform prevention and treatment efforts.

## Data Availability

Summary statistics from the GWAS will be available following publication through dbGaP at accession no. phs001672.v3.p 1

## References

1. Griswold MG, Fullman N, Hawley C, Arian N, Zimsen SRM, Tymeson HD, et al. Alcohol use and burden for 195 countries and territories, 1990–2016: a systematic analysis for the Global Burden of Disease Study 2016. The Lancet. 2018 sep 22;392(10152):1015–35.

2. American Psychiatric Association. Diagnostic and Statistical Manual of Mental Disorders [Internet]. Fifth Edition. American Psychiatric Association; 2013 [cited 2021 Jun 11]. Available from: http://psychiatryonline.org/doi/book/10.1176/appi.books.9780890425596

3. Clarke T-K, Adams MJ, Davies G, Howard DM, Hall LS, Padmanabhan S, et al. Genome-wide association study of alcohol consumption and genetic overlap with other health-related traits in UK Biobank (N=112 117). Mol Psychiatry. 2017 oct;22(10):1376–84.

4. Kranzler HR, Zhou H, Kember RL, Vickers Smith R, Justice AC, Damrauer S, et al. Genome-wide association study of alcohol consumption and use disorder in 274,424 individuals from multiple populations. Nat Commun. 2019 apr 2;10(1):1499.

5. Zhou H, Sealock JM, Sanchez-Roige S, Clarke T-K, Levey DF, Cheng Z, et al. Genome-wide meta-analysis of problematic alcohol use in 435,563 individuals yields insights into biology and relationships with other traits. Nat Neurosci. 2020 jul;23(7):809–18.

6. Bush K, Kivlahan DR, McDonell MB, Fihn SD, Bradley KA. The AUDIT alcohol consumption questions (AUDIT-C): an effective brief screening test for problem drinking. Ambulatory Care Quality Improvement Project (ACQUIP). Alcohol Use Disorders Identification Test. Arch Intern Med. 1998 sep 14;158(16):1789–95.

7. Saunders JB, Aasland OG, Babor TF, de la Fuente JR, Grant M. Development of the Alcohol Use Disorders Identification Test (AUDIT): WHO Collaborative Project on Early Detection of Persons with Harmful Alcohol Consumption--II. Addict Abingdon Engl. 1993 jun;88(6):791–804.

8. Sanchez-Roige S, Palmer AA, Fontanillas P, Elson SL, 23andMe Research Team, the Substance Use Disorder Working Group of the Psychiatric Genomics Consortium, Adams MJ, et al. Genome-wide association study meta-analysis of the Alcohol Use Disorders Identification Test (AUDIT) in two population-based cohorts. Am J Psychiatry. 2019 feb 1;176(2):107–18.

9. Liu M, Jiang Y, Wedow R, Li Y, Brazel DM, Chen F, et al. Association studies of up to 1.2 million individuals yield new insights into the genetic etiology of tobacco and alcohol use. Nat Genet. 2019 feb;51(2):237–44.

10. Sanchez-Roige S, Palmer AA, Clarke T-K. Recent efforts to dissect the genetic basis of alcohol use and abuse. Biol Psychiatry. 2020 apr 1;87(7):609–18.

11. Mallard TT, Savage JE, Johnson EC, Huang Y, Edwards AC, Hottenga JJ, et al. Item-level genome-wide association study of the Alcohol Use Disorders Identification Test in three population-based cohorts. Am J Psychiatry. 2021 may 14;appi.ajp.2020.20091390.

12. Dao C, Zhou H, Small A, Gordon KS, Li B, Kember RL, et al. The impact of removing former drinkers from genome-wide association studies of AUDIT-C. Addiction 2021 Apr 16. doi: 10.1111/add.15511. Online ahead of print.

13. Hatoum AS, Johnson EC, Polimanti R, Zhou H, Walters R, Consortium SUDWG of the PG, et al. The Addiction Genetic Factor a(g): A unitary genetic vulnerability characterizes substance use disorders and their associations with common correlates. medRxiv. 2021 jan 28;2021.01.26.21250498.

14. Gaziano JM, Concato J, Brophy M, Fiore L, Pyarajan S, Breeling J, et al. Million Veteran Program: A mega-biobank to study genetic influences on health and disease. J Clin Epidemiol. 2016 feb 1;70:214–23.

15. Chang CC, Chow CC, Tellier LC, Vattikuti S, Purcell SM, Lee JJ. Second-generation PLINK: rising to the challenge of larger and richer datasets. GigaScience [Internet]. 2015 Dec 1 [cited 2021 Jun 4];4(s13742-015-0047–8). Available from: https://doi.org/10.1186/s13742-015-0047-8

16. Willer CJ, Li Y, Abecasis GR. METAL: fast and efficient meta-analysis of genomewide association scans. Bioinforma Oxf Engl. 2010 sep 1;26(17):2190–1.

17. Yang J, Ferreira T, Morris AP, Medland SE, Madden PAF, Heath AC, et al. Conditional and joint multiple-SNP analysis of GWAS summary statistics identifies additional variants influencing complex traits. Nat Genet. 2012 apr;44(4):369–75.

18. Leeuw CA de, Mooij JM, Heskes T, Posthuma D. MAGMA: Generalized gene-get analysis of GWAS Data. PLOS Comput Biol. 2015 apr 17;11(4):e1004219.

19. Watanabe K, Taskesen E, van Bochoven A, Posthuma D. Functional mapping and annotation of genetic associations with FUMA. Nat Commun. 2017 nov 28;8(1):1826.

20. Bulik-Sullivan BK, Loh P-R, Finucane HK, Ripke S, Yang J, Patterson N, et al. LD Score regression distinguishes confounding from polygenicity in genome-wide association studies. Nat Genet. 2015 mar 1;47(3):291–5.

21. Brown BC, Asian Genetic Epidemiology Network Type 2 Diabetes Consortium, Ye CJ, Price AL, Zaitlen N. Transethnic genetic-correlation estimates from summary statistics. Am J Hum Genet. 2016 jul 7;99(1):76–88.

22. Baron RM, Kenny DA. The moderator–mediator variable distinction in social psychological research: Conceptual, strategic, and statistical considerations. J Pers Soc Psychol. 1986;51(6):1173–82.

23. Chang H, Xiao X, Li M. The schizophrenia risk gene ZNF804A : clinical associations, biological mechanisms and neuronal functions. Mol Psychiatry. 2017 jul;22(7):944–53.

24. Coleman JRI, Peyrot WJ, Purves KL, Davis KAS, Rayner C, Choi SW, et al. Genome-wide gene-environment analyses of major depressive disorder and reported lifetime traumatic experiences in UK Biobank. Mol Psychiatry. 2020 jul;25(7):1430–46.

25. Adewuyi EO, Mehta D, Sapkota Y, International Endogene Consortium, 23andMe Research Team, Auta A, et al. Genetic analysis of endometriosis and depression identifies shared loci and implicates causal links with gastric mucosa abnormality. Hum Genet. 2021 mar;140(3):529–52.

26. Hill WD, Marioni RE, Maghzian O, Ritchie SJ, Hagenaars SP, McIntosh AM, et al. A combined analysis of genetically correlated traits identifies 187 loci and a role for neurogenesis and myelination in intelligence. Mol Psychiatry. 2019 feb;24(2):169–81.

27. The GTEx Consortium. The GTEx Consortium atlas of genetic regulatory effects across human tissues. Science. 2020 sep 11;369(6509):1318–30.

28. Kang J, Coles M. IL-7: the global builder of the innate lymphoid network and beyond, one niche at a time. Semin Immunol. 2012 jun;24(3):190–7.

29. Nikou T, Ioannidis A, Zoga M, Tzavellas E, Paparrigopoulos T, Magana M, et al. Alteration in the concentrations of Interleukin-7 (IL-7), Interleukin-10 (IL-10) and Granulocyte Colony Stimulating Factor (G-CSF) in alcohol-dependent individuals without liver disease, during detoxification therapy. Drug Alcohol Depend. 2016 jun 1;163:77–83.

30. Lee JJ, Wedow R, Okbay A, Kong E, Maghzian O, Zacher M, et al. Gene discovery and polygenic prediction from a genome-wide association study of educational attainment in 1.1 million individuals. Nat Genet. 2018 jul 23;50(8):1112–21.

31. Morris AP, Voight BF, Teslovich TM, Ferreira T, Segrè AV, Steinthorsdottir V, et al. Large-scale association analysis provides insights into the genetic architecture and pathophysiology of type 2 diabetes. Nat Genet. 2012 sep;44(9):981–90.

32. de Vries PS, Brown MR, Bentley AR, Sung YJ, Winkler TW, Ntalla I, et al. Multiancestry Genome-Wide Association Study of Lipid Levels Incorporating Gene-Alcohol Interactions. Am J Epidemiol. 2019 jun 1;188(6):1033–54.

33. Edenberg HJ, McClintick JN. Alcohol dehydrogenases, aldehyde dehydrogenases, and alcohol use disorders: A critical review. Alcohol Clin Exp Res. 2018 dec;42(12):2281–97.

34. Buniello A, MacArthur JAL, Cerezo M, Harris LW, Hayhurst J, Malangone C, et al. The NHGRI-EBI GWAS Catalog of published genome-wide association studies, targeted arrays and summary statistics 2019. Nucleic Acids Res. 2019 jan 8;47(D1):D1005–12.

35. Speliotes EK, Willer CJ, Berndt SI, Monda KL, Thorleifsson G, Jackson AU, et al. Association analyses of 249,796 individuals reveal 18 new loci associated with body mass index. Nat Genet. 2010 nov;42(11):937–48.

36. Pardiñas AF, Holmans P, Pocklington AJ, Escott-Price V, Ripke S, Carrera N, et al. Common schizophrenia alleles are enriched in mutation-intolerant genes and in regions under strong background selection. Nat Genet. 2018 mar;50(3):381–9.

37. van der Meer D, Frei O, Kaufmann T, Shadrin AA, Devor A, Smeland OB, et al. Understanding the genetic determinants of the brain with MOSTest. Nat Commun. 2020 jul 14;11(1):3512.

38. Jones SE, Lane JM, Wood AR, van Hees VT, Tyrrell J, Beaumont RN, et al. Genome-wide association analyses of chronotype in 697,828 individuals provides insights into circadian rhythms. Nat Commun. 2019 jan 29;10(1):343.

39. Shrine N, Guyatt AL, Erzurumluoglu AM, Jackson VE, Hobbs BD, Melbourne CA, et al. New genetic signals for lung function highlight pathways and chronic obstructive pulmonary disease associations across multiple ancestries. Nat Genet. 2019 mar;51(3):481–93.

40. Said MA, van de Vegte YJ, Verweij N, van der Harst P. Associations of observational and genetically determined caffeine intake with coronary artery disease and diabetes mellitus. J Am Heart Assoc. 2020 dec 15;9(24):e016808.

