## Supplemental Methods for "Genetic underpinnings of the transition from alcohol consumption to alcohol use disorder: shared and unique genetic architectures in a cross-ancestry sample"

*Million Veteran Program Cohort*

Recruitment for MVP began in 2011 and is ongoing, with approximately 830,000 participants enrolled at 63 VA medical centers nationwide as of June 2021. Written informed consent was obtained from all MVP study participants. Following consent, Veterans provided a blood sample for DNA extraction and genotyping, many completed surveys about health and lifestyle, and all allowed access to their EHR for research purposes. The MVP received approval from the Central Veterans Affairs Institutional Review Board (IRB) and site-specific IRBs. All relevant ethical regulations for work with human subjects were followed in the conduct of the study.

*Phenotypes*

*Primary GWAS*: For the primary AUD GWAS (“AUD [Stringent]”), the case definition required the presence of at least one inpatient or two outpatient ICD-9/10 diagnostic codes for AUD (305.0X, 303.X, F10.1X, F10.2X) from 2000-2018, consistent with our previous work^1,2^. This approach is generally preferred when using diagnostic codes in the outpatient setting because these codes are sometimes incorrectly used in the process of ruling out the condition rather than establishing the condition. Thus, individuals with only one outpatient diagnostic code for AUD were excluded from the analysis. Controls were individuals who had no AUD ICD-9/10 diagnostic code in the EHR. For the primary AUDIT-C GWAS, maximum AUDIT-C score was used as a measure of alcohol consumption (“AUDIT-C”).

*Secondary GWAS*: For the secondary AUD GWASs we created 1) a less stringent definition of AUD (“AUD [Less stringent]”) that required just one inpatient or outpatient ICD-9/10 diagnostic code for AUD in the EHR, and 2) removed cases and controls with a maximum AUDIT-C=0 (thus, all individuals remaining in the analysis reported alcohol consumption; “AUD [AUDIT-C>0]”). Controls were individuals who had no AUD ICD-9/10 diagnostic code in the EHR. For the secondary AUDIT-C GWAS, we removed individuals with a maximum AUDIT-C=0 ( “AUDIT-C>0”), to avoid the confounding effects of a prior history of heavy drinking despite current abstinence^3^.

*Mediation analyses*: For mediation analyses, cases were defined as those meeting the stringent definition for AUD. As in previous analyses, controls were individuals who did not have an AUD ICD-9/10 diagnostic code in the EHR. We excluded abstainers (individuals with AUDIT-C score=0) from the analysis because this group is heterogenous, consisting of both never- and former drinkers, of which the latter group could give rise to reverse causal effects. For AUD cases, we required that the AUDIT-C score pre-date the AUD diagnosis to minimize the possibility of reverse causality. Therefore, for AUD cases we used maximum AUDIT-C score prior to the date of the first ICD-9/10 AUD code as their exposure variable. For controls, we used the maximum AUDIT-C score ever measured. If either cases or controls had multiple instances of the maximum AUDIT-C score, the first instance was used.

*Genotyping and imputation*

Genotyping was performed using a custom Affymetrix Axiom Biobank Array. Genotyping data for MVP have been released in batches as recruitment is ongoing; analyses presented in this paper used MVP Release 3 data. Quality control was performed by the MVP Genomics working group prior to imputation. Samples with excessive heterozygosity or missing call rate > 2.5% or variants with low call rate or deviation from expected allele frequency were removed, leaving 455,789 individuals and 668,280 markers.

Genotypes were phased and imputed with EAGLE v2^4^ and Minimac4^5^, with the 1000 Genomes Project phase 3, version 5^6^ reference panel. Principal components (PCs) were computed using FlashPCA^7^ on all MVP participants and 2,504 individuals from 1000 Genomes. These were used to infer genetic ancestry, which was unified with self-identified race/ethnicity using the HARE (Harmonizing Genetic Ancestry and Self-identified Race/Ethnicity) method ^8^ to construct ancestry groups. A total of 87,169 African Americans (AAs); 318,725 European Americans (EAs); and 34,160 Hispanic American (HAs) were identified. We did not include individuals from other population groups due to their small numbers.

Quality control of variants and individuals was conducted within each ancestry group. We excluded one individual randomly from each pair of related individuals (kinship coefficient=0.0884). Variants were excluded based on minor allele frequency (AA < 0.001; EA < 0.0005; HA < 0.005), genotype call rate ≤ 0.95, and Hardy-Weinberg equilibrium p-value ≤ 1x10^-6^. Population-specific imputation INFO scores were calculated using SNPTEST v2^9^ and variants with INFO scores < 0.3 were removed. The number of variants remaining was approximately 19.5 million in AAs, 11.8 million in EAs, and 8.8 million in HAs.

*Independent variants*

To identify independent variants in a region, we performed LD-clumping using a range of 3000 kb and r^2^ > 0.1, with the matched 1000 Genomes reference panel as the background. If independent variants were located < 1Mb apart they were merged into a single locus. Due to the known long-range LD in the ADH gene cluster on chromosome 4, all variants within chr4q23-q24 (~97.2-102.6 Mb) were merged into a single locus. If a locus contained multiple variants, we conducted conditional analyses using GCTA-COJO^10^ to define conditionally independent variants. For each locus, the most significant (index) variant was conditioned on, using the matched ancestry subjects from 1000 Genomes as the LD reference sample. Variants that remained significant following conditioning (p < 5x10^-8^) were subject to another round of conditional analyses by including the next most significant variant in an iterative process until all independent variants were identified.

*Heritability analyses and genetic correlation*

Population-specific LD scores were calculated based on the 1000 Genomes Phase 3 dataset for African and European ancestries, limited to HapMap3^11^ variants following removal of the major histocompatibility complex region (chr6:26-34 Mb). LD scores were calculated for 226,116 variants in AAs and 399,040 variants in EAs. LD scores were not calculated for the HA group due to the smaller sample size. Summary statistics for each phenotype were limited to HapMap3 variants and used to estimate h^2^_SNP_. Genetic correlation (r_g_) between phenotypes within each ancestry was estimated using LDSC and the corresponding 1000 Genomes population as the reference panel. Cross-ancestry genetic correlations between AAs and EAs within phenotypes were estimated using POPCORN^12^. Genetic correlations with 257 published traits were estimated using LD Hub^13^ and the EA summary statistics for each phenotype. Correlations were considered significant using a Bonferroni-corrected p-value (1.95x10^-3^).

1.	Kranzler, H. R. *et al.* Genome-wide association study of alcohol consumption and use disorder in 274,424 individuals from multiple populations. *Nat. Commun.* **10**, 1499 (2019).

2.	Zhou, H. *et al.* Genome-wide meta-analysis of problematic alcohol use in 435,563 individuals yields insights into biology and relationships with other traits. *Nat. Neurosci.* **23**, 809–818 (2020).

3.	Dao, C. *et al.* The impact of removing former drinkers from genome-wide association studies of AUDIT-C. *Addiction* **n/a**,.

4.	Loh, P.-R. *et al.* Reference-based phasing using the Haplotype Reference Consortium panel. *Nat. Genet.* **48**, 1443–1448 (2016).

5.	Das, S. *et al.* Next-generation genotype imputation service and methods. *Nat. Genet.* **48**, 1284–1287 (2016).

6.	Auton, A. *et al.* A global reference for human genetic variation. *Nature* **526**, 68–74 (2015).

7.	Abraham, G., Qiu, Y. & Inouye, M. FlashPCA2: principal component analysis of Biobank-scale genotype datasets. *Bioinformatics* **33**, 2776–2778 (2017).

8.	Fang, H. *et al.* Harmonizing Genetic Ancestry and Self-identified Race/Ethnicity in Genome-wide Association Studies. *Am. J. Hum. Genet.* **105**, 763–772 (2019).

9.	Marchini, J. & Howie, B. Genotype imputation for genome-wide association studies. *Nat. Rev. Genet.* **11**, 499–511 (2010).

10.	Yang, J. *et al.* Conditional and joint multiple-SNP analysis of GWAS summary statistics identifies additional variants influencing complex traits. *Nat. Genet.* **44**, 369–375 (2012).

11.	Altshuler, D. M. *et al.* Integrating common and rare genetic variation in diverse human populations. *Nature* **467**, 52–58 (2010).

12.	Brown, B. C., Asian Genetic Epidemiology Network Type 2 Diabetes Consortium, Ye, C. J., Price, A. L. & Zaitlen, N. Transethnic Genetic-Correlation Estimates from Summary Statistics. *Am. J. Hum. Genet.* **99**, 76–88 (2016).

13.	Zheng, J. *et al.* LD Hub: a centralized database and web interface to perform LD score regression that maximizes the potential of summary level GWAS data for SNP heritability and genetic correlation analysis. *Bioinformatics* **33**, 272–279 (2017).
