## Supplemental Figures for "Genetic underpinnings of the transition from alcohol consumption to alcohol use disorder: shared and unique genetic architectures in a cross-ancestry sample"

### Supplemental Figure 1: Analytic methods

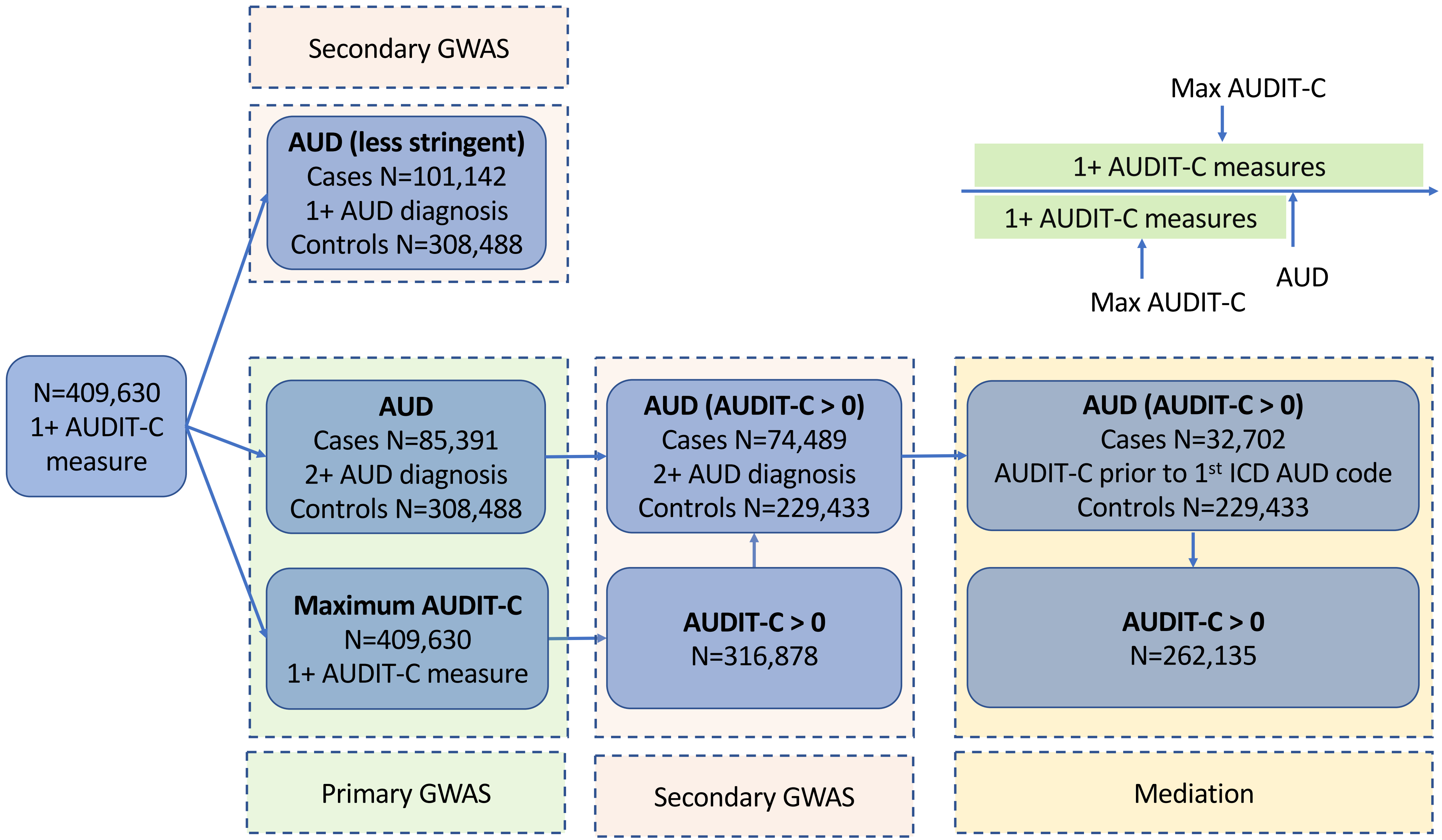

### Supplemental Figure 2: AUDIT-C scores

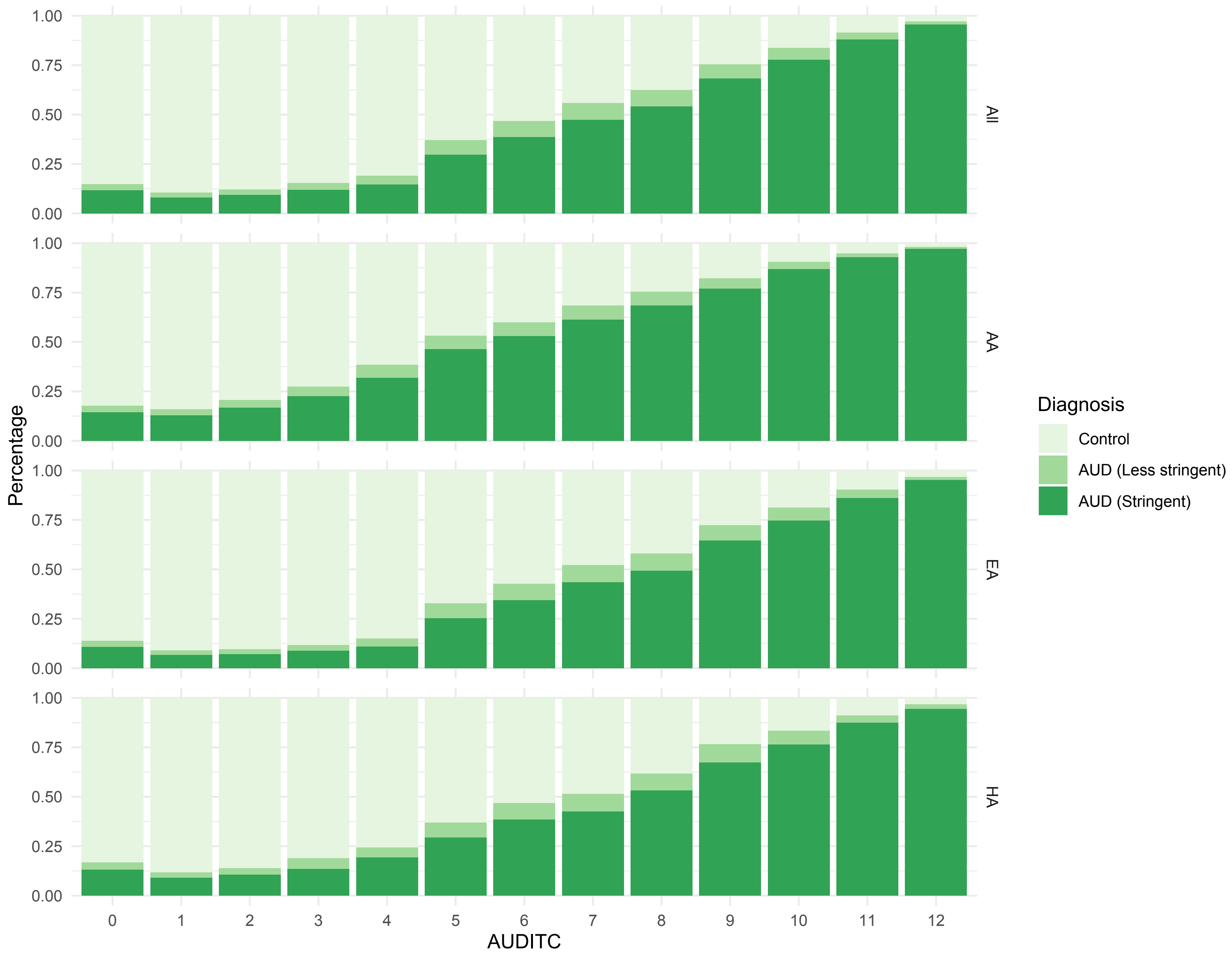

### Supplemental Figure 3: Summary of GWAS results

Numbers in circles at top represent number of GWS loci overlapping between GWAS

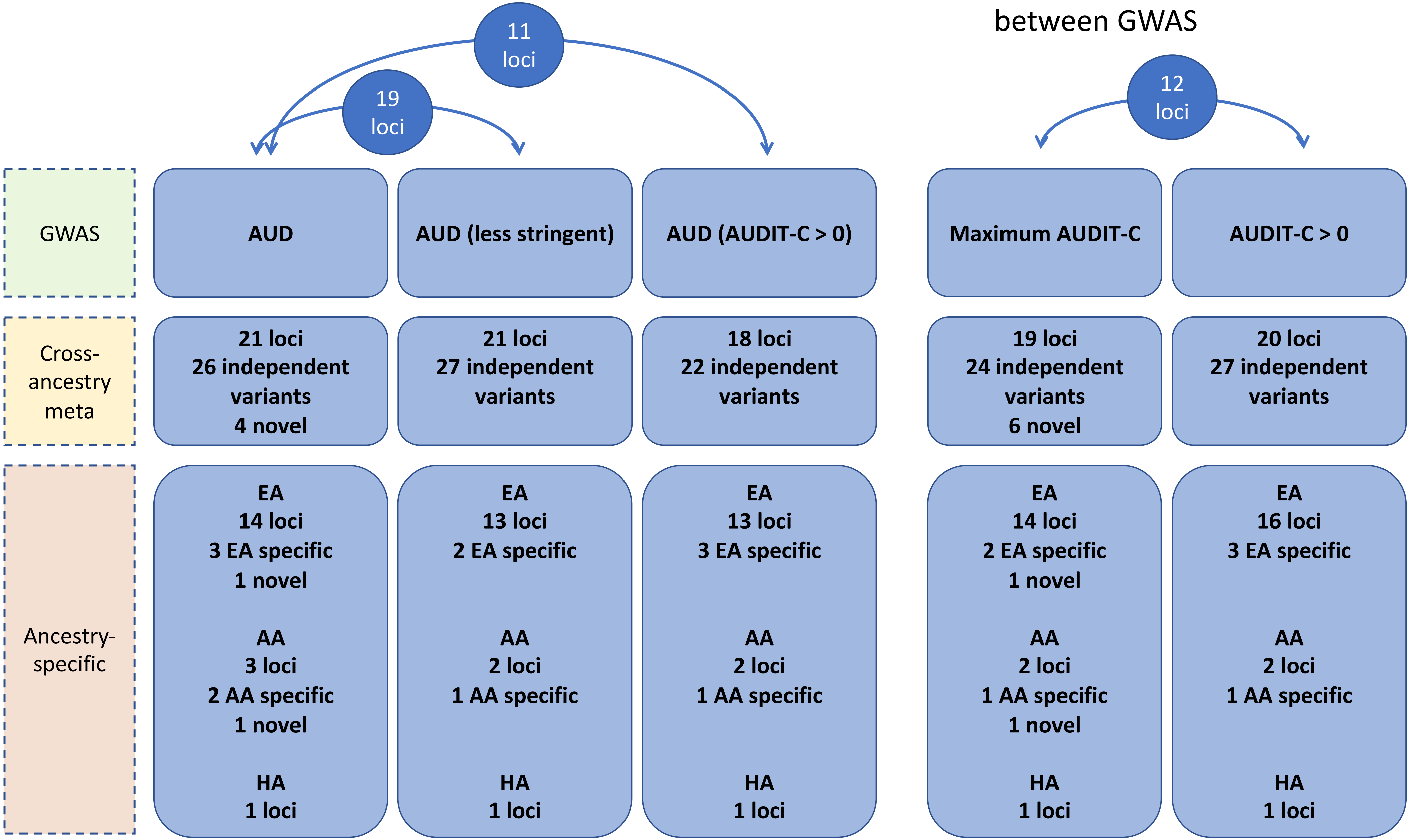

Supplemental Figure 4: Manhattan plots for AUD by ancestry

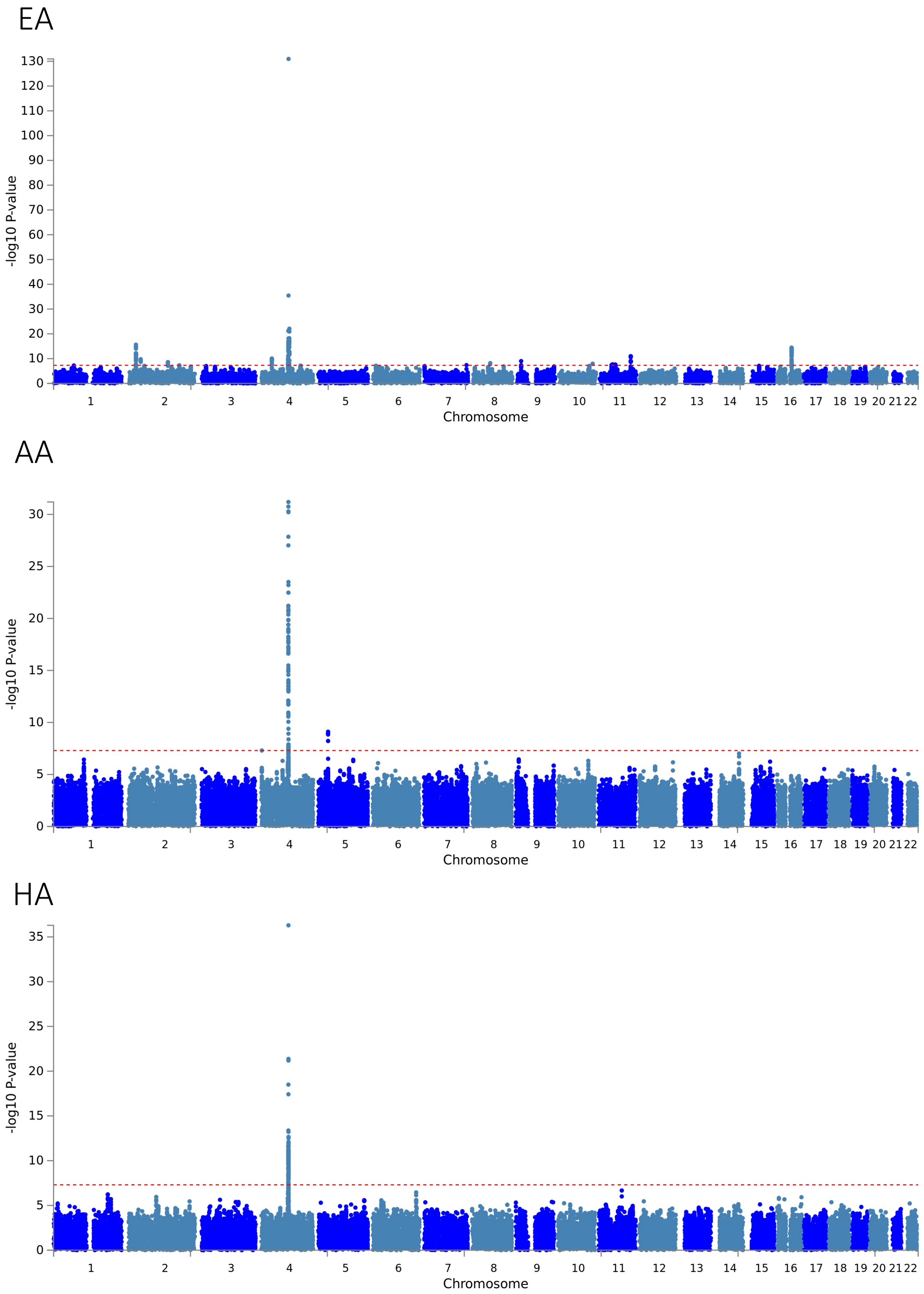

Supplemental Figure 5: Locus zoom plots for ancestry-specific variants associated with AUD in EAs

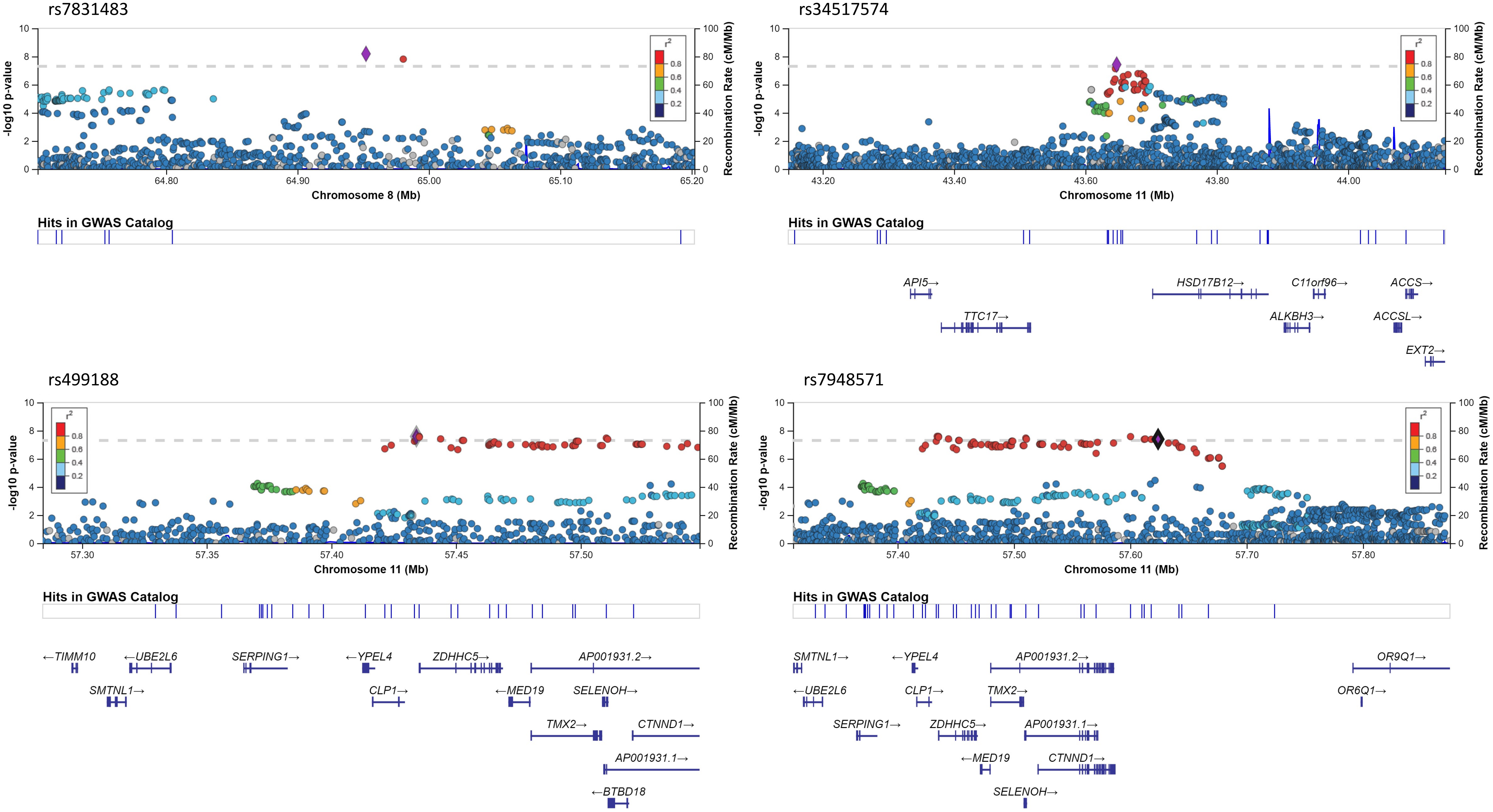

### Supplemental Figure 6: Locus zoom plots for ancestry-specific variants associated with AUD in AAs

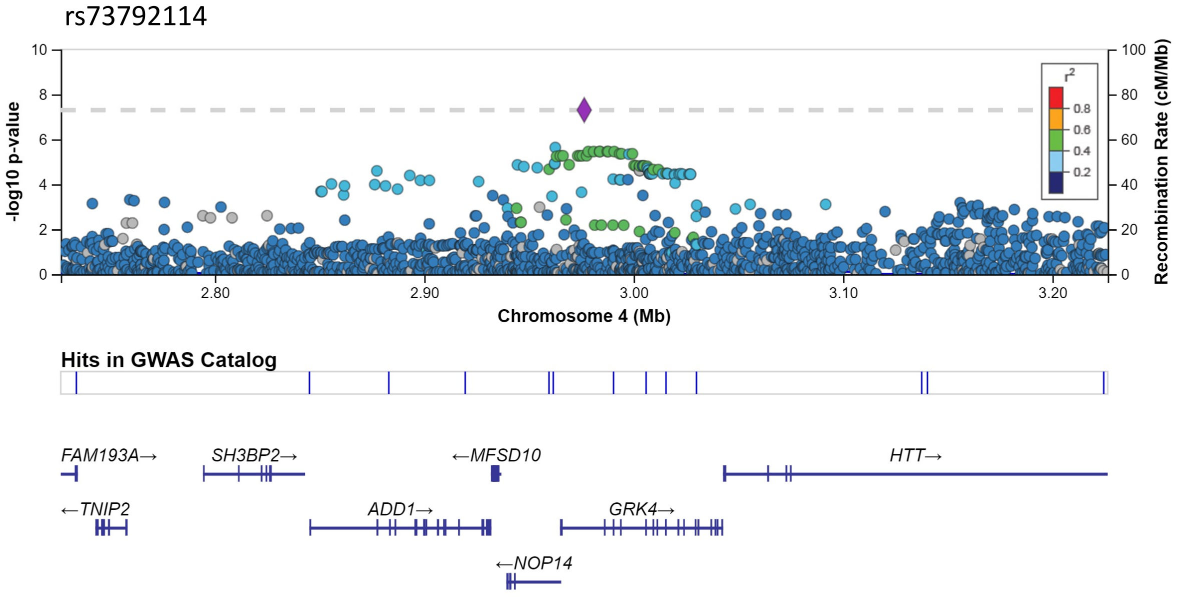


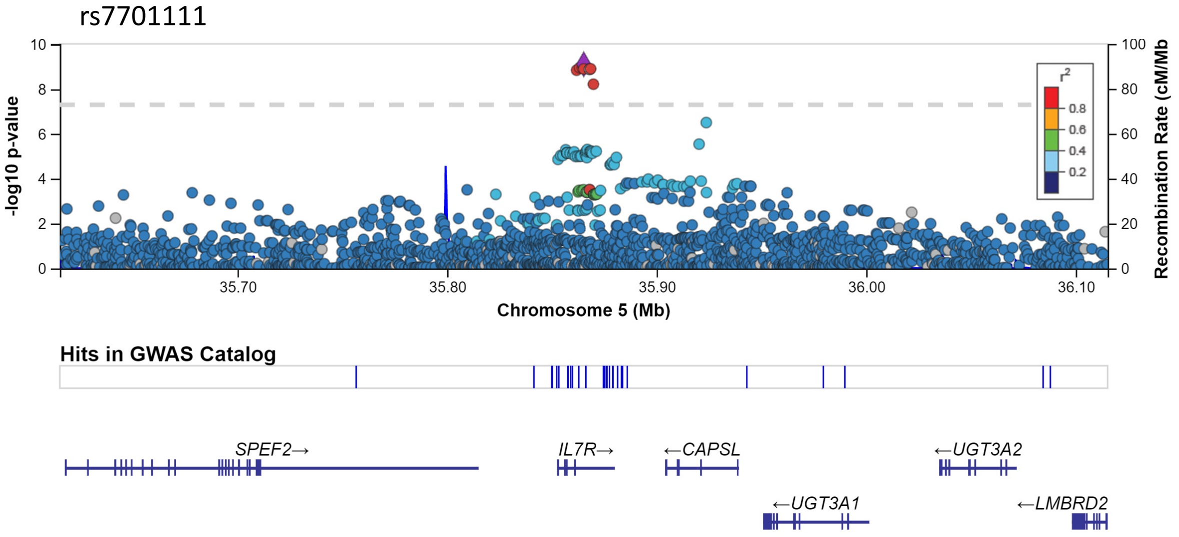

Supplemental Figure 7: Manhattan plots for AUDIT-C by ancestry

EA

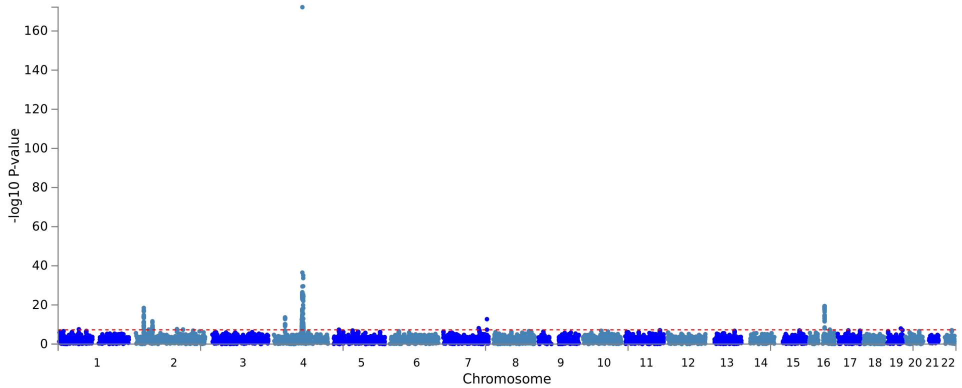


AA

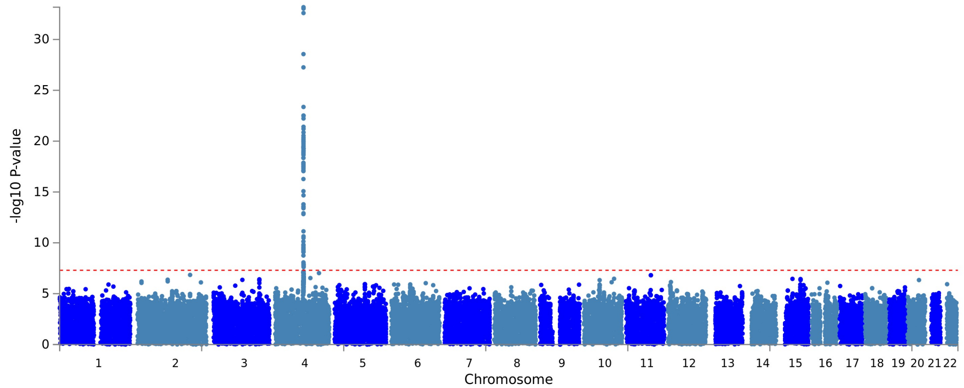


HA

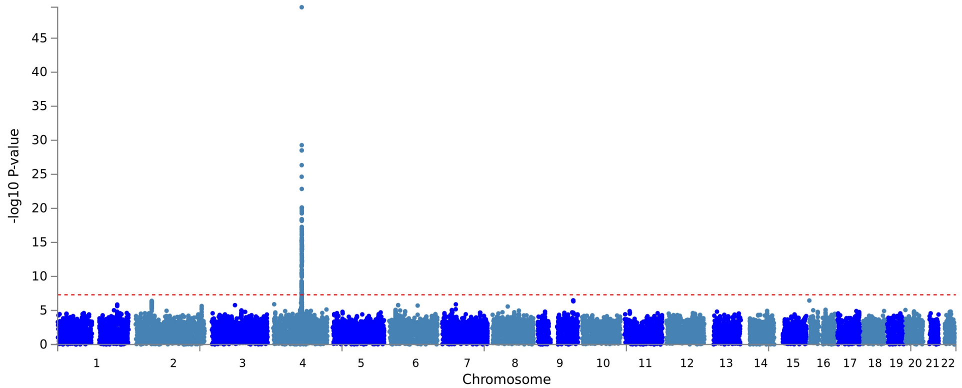

### Supplemental Figure 8: Locus zoom plots for ancestry-specific variants associated with AUDIT-C in EAs

rs13389219

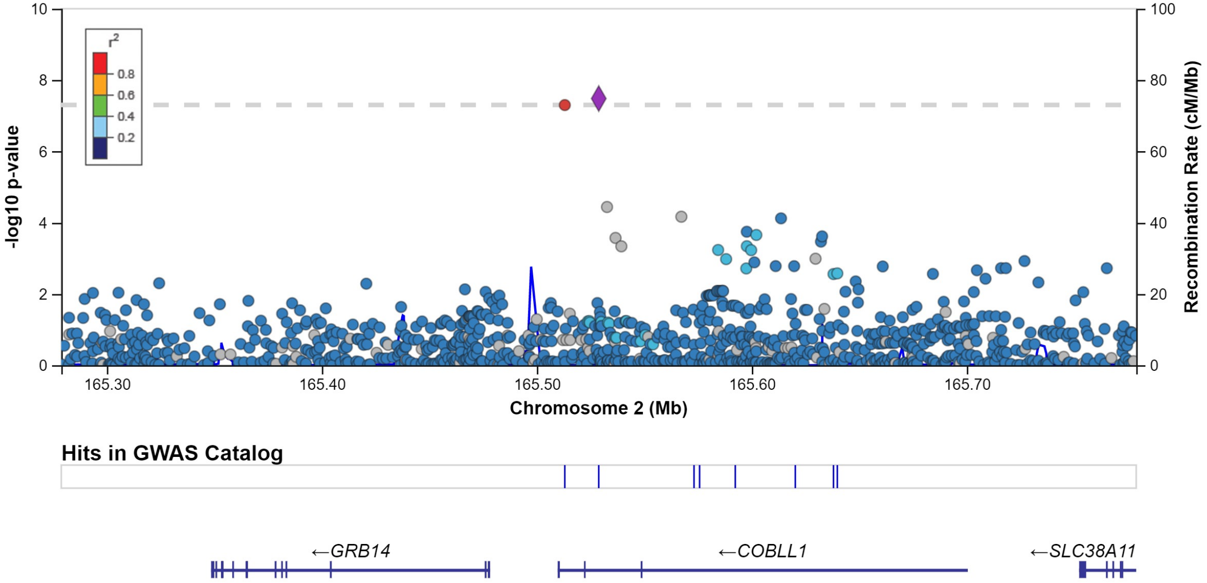


rs72740892

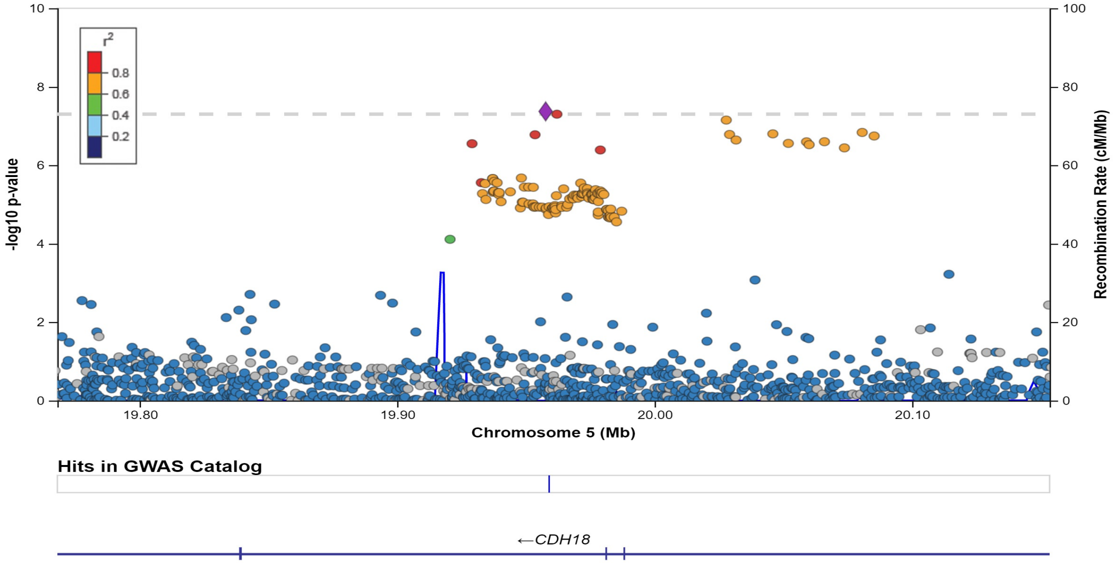

### Supplemental Figure 9: Locus zoom plots for ancestry-specific variants associated with AUDIT-C in AAs

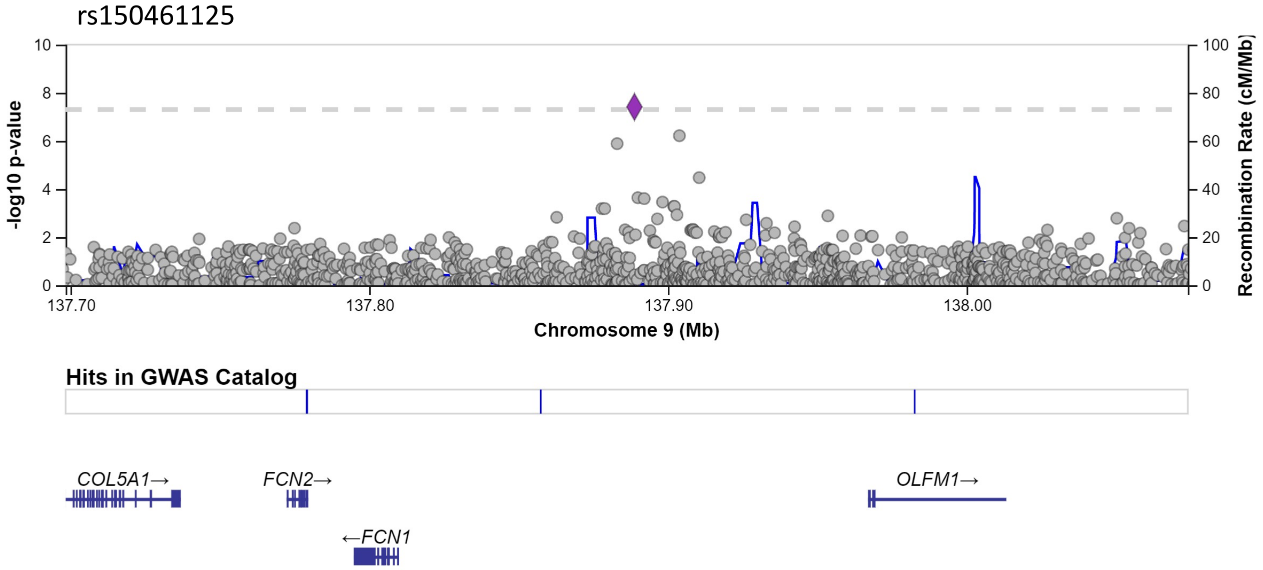

### Supplemental Figure 10: Manhattan plot for AUD (Less Stringent)

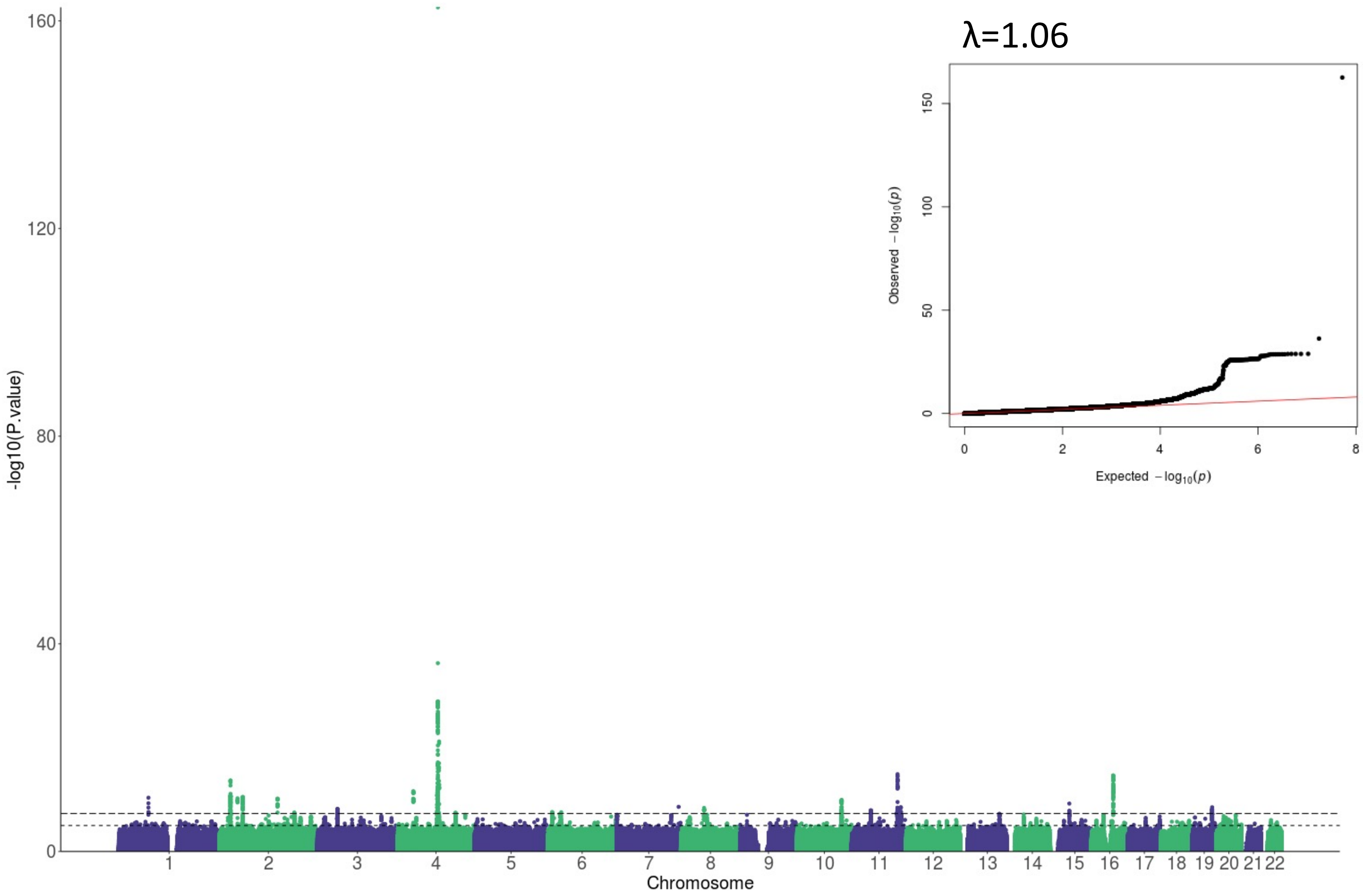

### Supplemental Figure 11: Manhattan plot for AUD (Stringent, AUDIT-C>0)

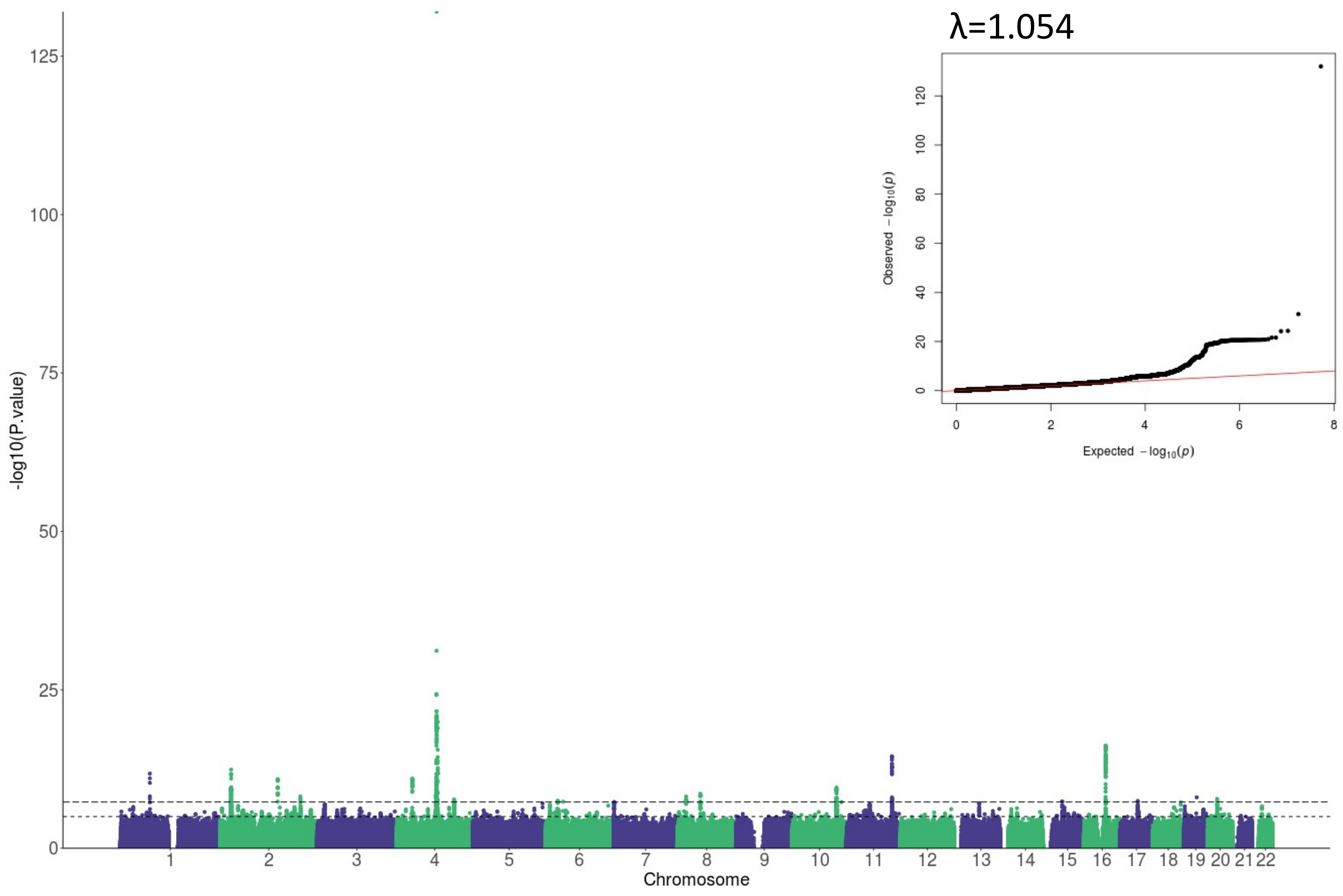

### Supplemental Figure 12: Manhattan plot for AUDIT-C>0

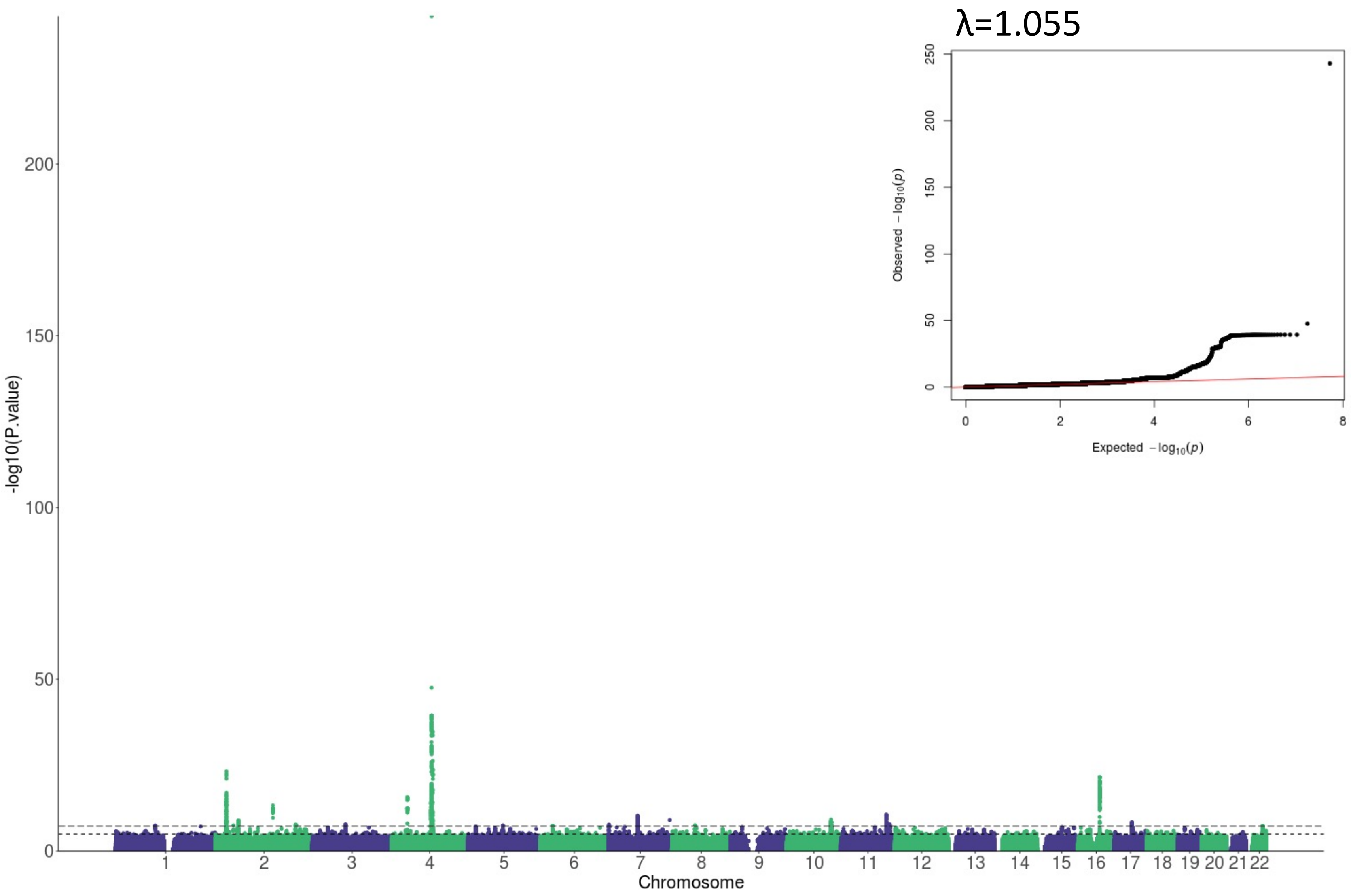

### Supplemental Figure 13: Gene-based association analyses for AUD

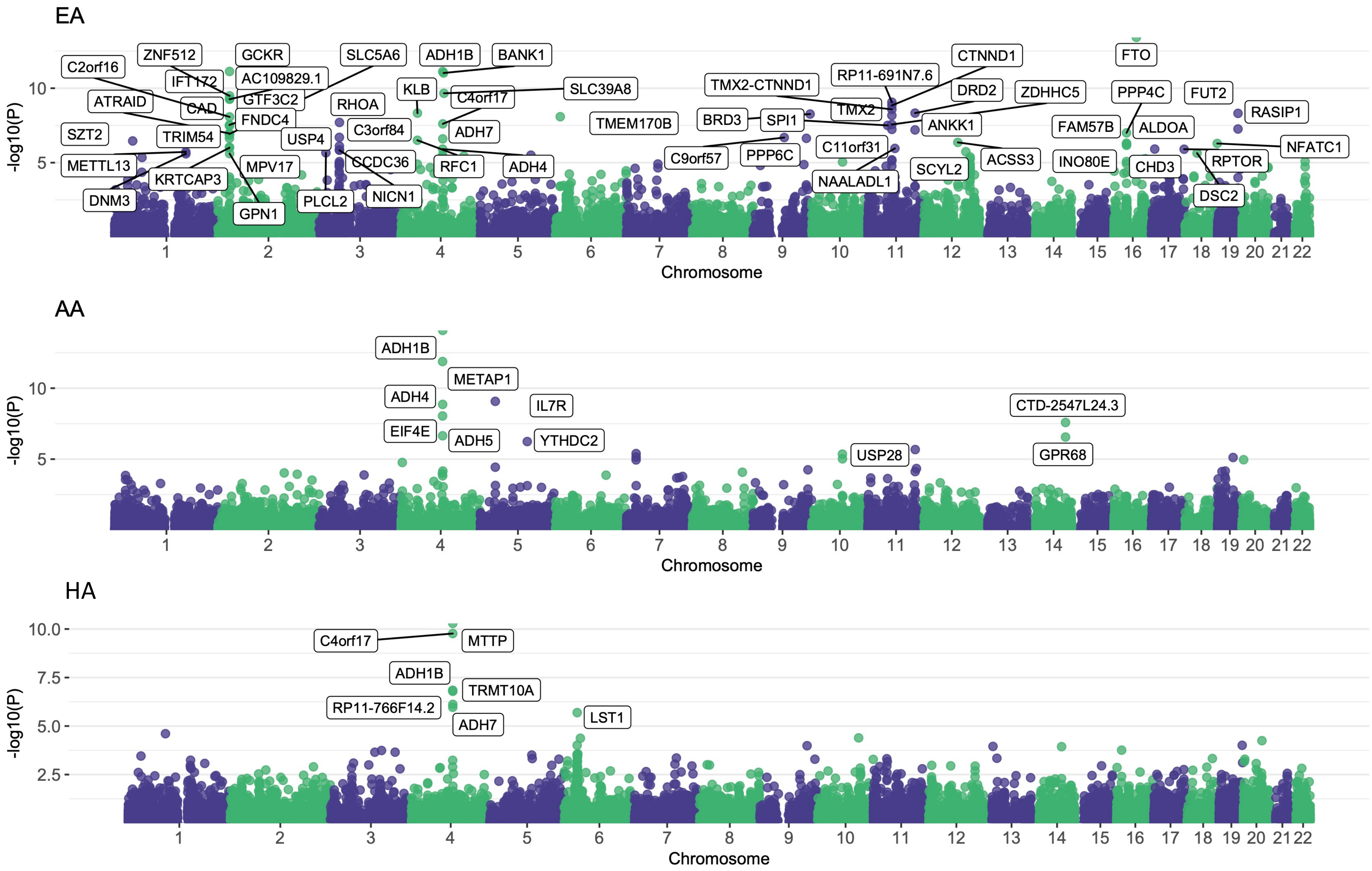

### Supplemental Figure 14: Gene-based association analyses for AUDIT-C

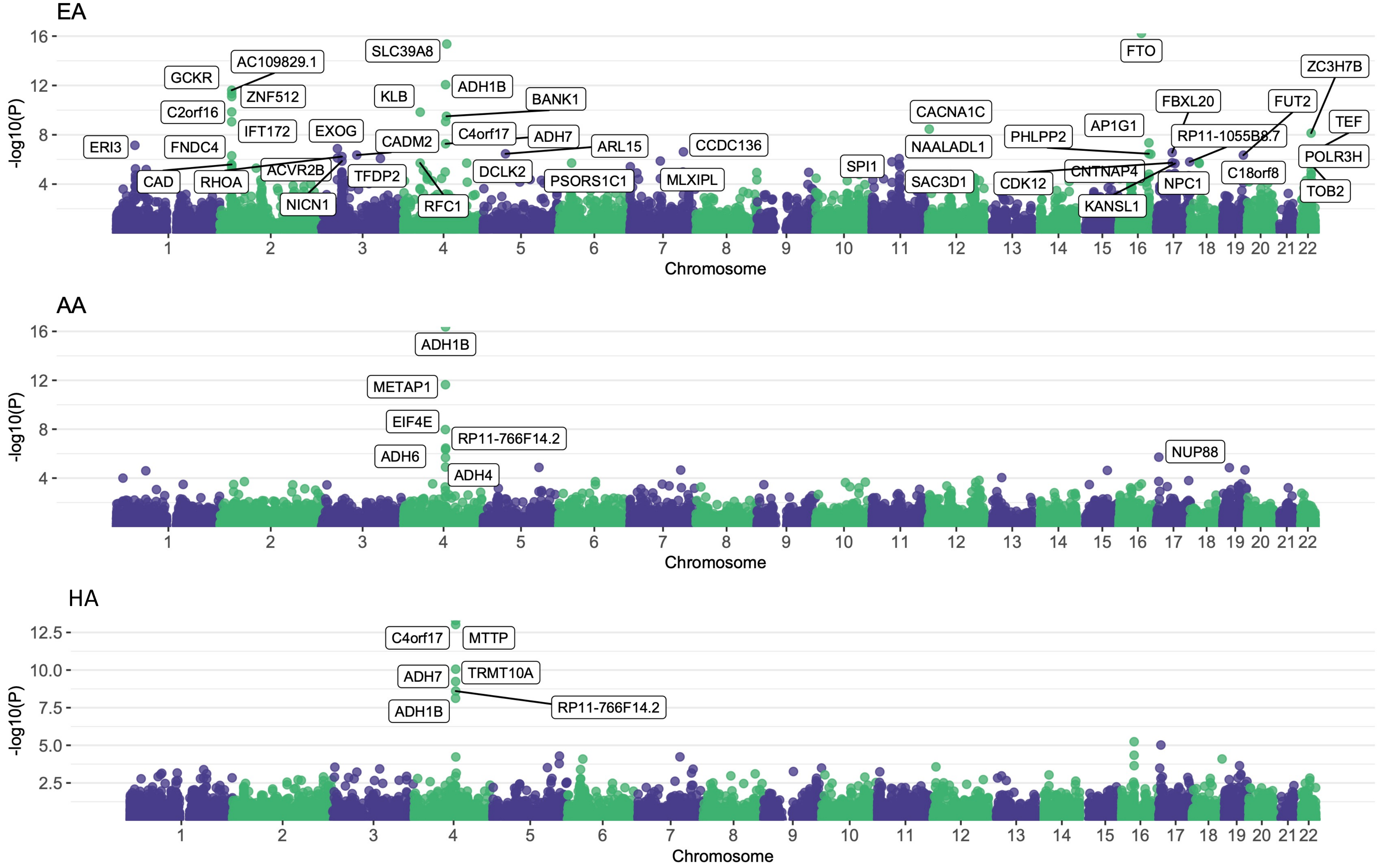

### Supplemental Figure 15: Genetic correlation between phenotypes

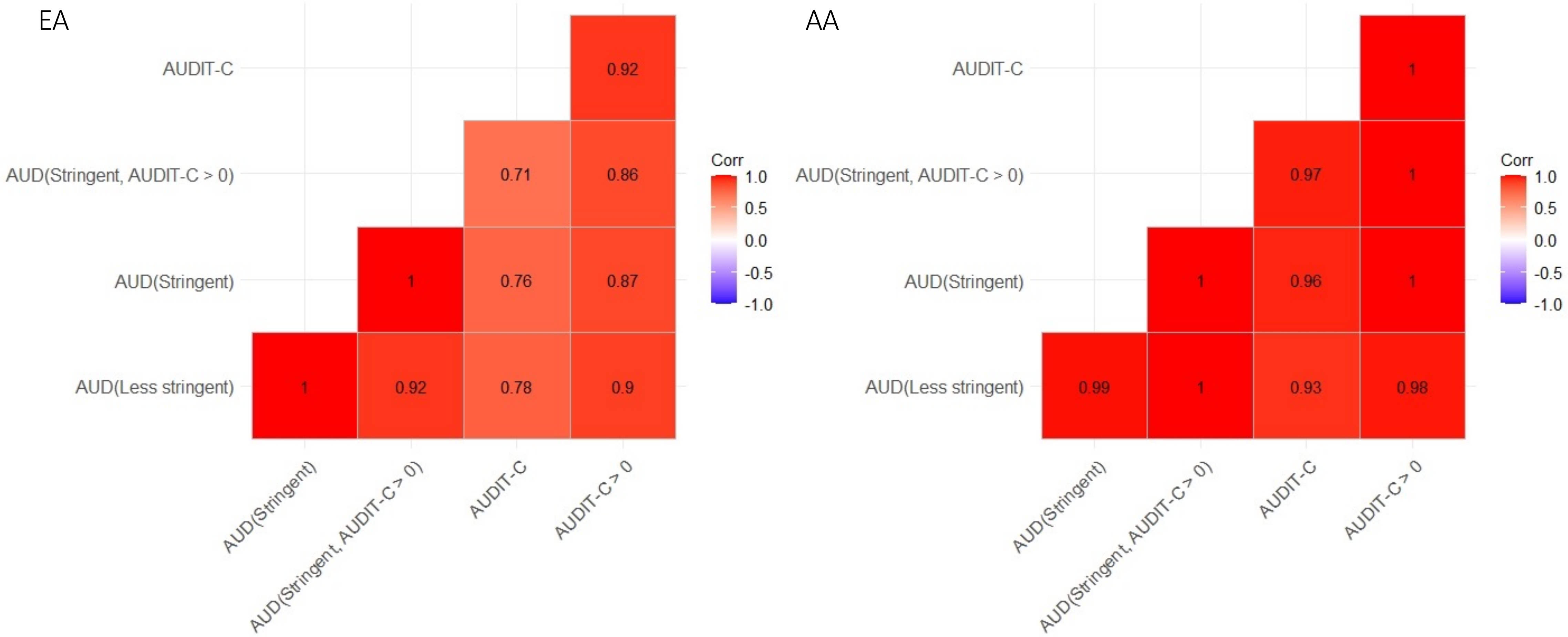
